# Glial activation mediates phenotypic effects of *APOEε4* and sex in Alzheimer’s disease

**DOI:** 10.1101/2024.03.08.24303882

**Authors:** Roger M. Lane, Dan Li, Taher Darreh-Shori

**Affiliations:** Ionis Pharmaceuticals Inc., Carlsbad, CA, USA; Karolinska Institutet, Department of Neurobiology, Care Sciences and Society; Division of Clinical Geriatrics, Stockholm, Sweden

**Keywords:** Apolipoprotein E, sex, glial activation, amyloid, tau, synaptic injury

## Abstract

**INTRODUCTION:** This study examined the impact of *apolipoprotein ɛ4* (*APOEɛ4*) allele frequency and sex on the phenotype of Alzheimer’s disease (AD).

**METHODS:** The baseline characteristics, CSF, and neuroimaging biomarkers, and cognition scores collected from 45 patients aged 50-74 years with confirmed early AD from clinical trial NCT03186989 were evaluated in a post-hoc study.

**RESULTS:** A phenotypic spectrum was observed from a predominant amyloid and limbic-amnestic phenotype in male *APOEɛ4* homozygotes to a predominantly tau, limbic-sparing, and multidomain cognitive impairment phenotype in female *APOEɛ4* noncarriers. Amyloid pathology inversely correlated with tau pathophysiology, glial activation, and synaptic injury, with the strongest correlations observed in male *APOEɛ4* carriers. Tau pathophysiology was correlated with glial activation, synaptic injury, and neuroaxonal damage, with the strongest correlation observed in female *APOEɛ4* noncarriers.

**DISCUSSION:** Glial activation is influenced by apoE isoform and sex, which explains much of the phenotypic heterogeneity in early AD below age 75 years.

**HIGHLIGHTS:**

- *APOEɛ4* homozygotes displayed a predominantly amyloid and limbic-amnestic phenotype.
- Female *APOEɛ4* noncarriers displayed a predominantly tau, limbic-sparing, and multidomain cognitive impairment phenotype.
- In male *APOEɛ4* carriers, amyloid pathology was inversely correlated with tau pathophysiology, synaptic injury, and glial activation
- Females displayed a non-*APOEɛ4* allele frequency-dependent increase in glial activation and synaptic injury
- In female *APOEɛ4* noncarriers, tau pathophysiology was strongly correlated with glial activation, synaptic injury, and neuroaxonal damage

**RESEARCH IN CONTEXT:** *Systematic review:* The impact of *APOEɛ4* alleles and sex on phenotypic features was examined in 45 patients, aged 50-74 years, with early AD.

*Interpretation:* Findings were consistent with prior reports and suggest that glial activation, influenced by apoE isoform and sex, explains much of the phenotypic heterogeneity in early AD below age 75 years. Lower glial activation in *APOEɛ4* homozygotes associated with the highest levels of amyloid and the lowest levels of tau pathology, and a limbic-amnestic phenotype, suggesting degeneration of basal forebrain cholinergic neurons. Higher glial activation in female *APOEɛ4* noncarriers was associated with the highest tau pathology and synaptic injury, the lowest amyloid pathology, greater ventricular expansion, and multi-domain cognitive deficits.

*Future directions:* This work defined a combined sex, genotype, and age framework that delineates multiple pathways to end-stage AD. Confirmation is required, followed by optimization of therapeutic approaches to amyloid, tau, and glial activation pathologies along the disease stage continuum.

## 1. BACKGROUND

The *APOEɛ4* polymorphism is the major genetic risk factor for sporadic AD, mainly exerting its pathogenic effects via altered glial activation and amyloid pathology accumulation (1, 2). Functional levels of glial activation may balance the cellular needs to stimulate amyloid-beta (Aβ) clearance and limit both synaptic engulfment and the spread of tau pathology (3–6). Glial activation is associated with the spread of tau and synaptic pathology (5, 7, 8). In *APOEɛ4* noncarriers, females have higher levels of CSF tau pathology in both prodromal disease and mild AD compared to males (9). Female noncarriers are also more likely to develop overactivated microglia with age (10). In females with AD, glial activation is generally higher (11) and cortical microglial activation is more important for AD progression (12). In contrast, glial responses in *APOEɛ4* carriers are deficient and favor the accumulation of amyloid pathology (13). This deficiency implicates altered lipid dynamics, altered cholinergic and checkpoint signaling, and may also be a consequence of Aβ accumulation.

Poor lipidation of apoE4 impairs its ability to opsonize extracellular debris such as fibrillar Aβ_42_ and myelin breakdown products, which impairs microglial recognition and clearance responses (14, 15). Rather than clear debris effectively, *APOEɛ4* microglia show phagocytic and endolysosomal dysfunction, become senescent, upregulate pro-inflammatory response genes, and downregulate genes responsible for migration and phagocytosis (16, 17). Like overactive glia, these functionally underactive glia produce proinflammatory cytokines (18), and therefore, it is critical that biomarkers of glial activation represent functional activation of glia and not inflammation (19).

ApoE4 is associated with decreased intracellular exchange of fatty acids and cholesterol between neurons and astrocytes with reduced fatty acid β-oxidation, lipid droplet accumulation, increased neuronal cholesterol synthesis, and impaired abilities to maintain myelin and cell membranes (20). Fatty-acid fueled bioenergetics improve microglial migration, phagocytic ability, and lysosomal degradation (21, 22), whereas the loss of these functions is associated with lipid droplet formation that is markedly exacerbated in *APOEɛ4* carriers and by proximity to Aβ plaques (23). Altered lipid composition and fluidity of the cell membrane alters the stability and function of membrane receptors, including alpha 7 nicotinic acetylcholine (ACh) receptors (α7-nAChRs)(24, 25).

The cholinergic system acts to control glial reactivity and function via rapid focal synaptic signaling and slow diffuse extracellular signaling though α7-nAChRs (26). In a concentration and aggregation-dependent manner, Aβ signals through α7-nAChRs and also influences the extracellular fluid equilibrium between the breakdown and synthesis of ACh via effects on ACh-hydrolyzing capacity of cholinesterases (27) and choline acetyltransferase (ChAT) activity (28). ApoE forms soluble and highly stable complexes with cholinesterase enzymes and Aβ, that can oscillate between slow and ultrafast ACh hydrolysis, depending on Aβ availability (29). Reduced cholinesterase activity and lowering of glial activation markers are seen in *APOEɛ4* carriers, particularly in individuals with polymorphic variants of genes encoding cholinesterase enzymes with lower activity (29, 30). These individuals accumulate Aβ earlier with a younger age-of-onset of AD (31). Amnestic symptoms parallel the progressive denervation of basal forebrain corticolimbic cholinergic neuronal projections that remove the cholinergic ‘brake’ on glial activation in the medial temporal lobe (MTL) and other cortical regions, permitting the spread of tau and neurodegenerative pathology, and transitioning these individuals into AD (31, 32).

This study sought to examine the early AD phenotypic spectrum with respect to *APOEɛ4* genotype. The study analyzed cross-sectional demographic, CSF biomarker, neuroimaging and cognitive domain assessments from patients below 75 years of age who were diagnosed with early AD. The younger age of participants limited the likelihood of other age-related pathologies, which likely allowed for a clearer examination of the influence of genotype on early AD.

## 2. METHODS

### 2.1 Clinical Trial

Baseline samples obtained from Clinical Trial NCT03186989 were assessed for biomarkers in a retrospective study. The trial was conducted at 12 centers in Canada, Finland, Germany, the Netherlands, Sweden, and the UK between August 2017 and February 2020. The trial was conducted in accordance with Good Clinical Practice Guidelines of the International Council for Harmonisation, and according to the ethical principles outlined in the Declaration of Helsinki. The complete study (33) was approved by relevant ethics committees (**Table S1**). Written informed consent was provided by the participants.

### 2.2 Study Eligibility

Eligible study participants were between the ages of 50 and 74; had probable early AD (amnestic or non-amnestic), defined by a Mini-Mental State Examination score (MMSE) (34) of 20-27, inclusive, and either a Clinical Dementia Rating (35) Overall Global Score of 1, or a Global Score of 0.5 with a Memory Score of 1; a CSF pattern of low Aβ_1-42_ (≤ 1200 pg/ml), elevated total-tau (> 200 pg/ml) and p-tau (> 18 pg/ml), and a total-tau to Aβ_1-42_ ratio > 0.28; and a diagnosis of probable AD based on National Institute of Aging-Alzheimer’s Association (NIA-AA) criteria (36).

### 2.3 Data Collection and Processing

CSF collected from participants at the baseline of an interventional study was analyzed for markers of amyloid accumulation (inversely indexed by Aβ_42_), tau pathophysiology (p-tau_181_), neuroaxonal degeneration (NfL), synaptic injury (Ng), glial activation (YKL-40) (**Table S2**). Patients were characterized for common variants of *APOE* (ie, *APOEɛ4*, *APOEɛ3*, and *APOEɛ2*), and butyrylcholinesterase *(BCHE)* (ie, *BCHE-K*).

The study required 3-dimensional T1-weighted structural magnetic resonance imaging (MRI) scans of the head with volumetric analyses calculated using VivoQuant^TM^, which includes preprocessing and multi-atlas segmentation modules, followed by visual inspection and manual editing if needed (37). The mean baseline ventricular volume and hippocampal volume were expressed as a percentage of the total intracranial volume (% ICV). Baseline Mini-Mental State Examination (MMSE) and Repeatable Battery for the Assessment of Neurological Status (RBANS) scores were also reported.

### 2.4 Statistical analysis

Quantitative assessments were summarized using descriptive statistics, including number of patients, mean, and standard deviation. Qualitative assessments were summarized using frequency counts and percentages. The exact test was used to examine the Hardy-Weinberg equilibrium (HWE) in the distribution of *APOEɛ4* alleles in the study population. All exact tests were performed using the R package “Hardy Weinberg” (38).

Analysis of variance (ANOVA) and analysis of covariance (ANCOVA) were used to test whether the burden of amyloid pathology differed across *APOEɛ4* homozygotes, heterozygotes, and noncarriers. When the ANCOVA model was applied, the model included *BCHE-K* carrier status and sex as factors and the baseline MMSE total score as a covariate. Prior to performing ANOVA and ANCOVA, the normality assumption of residuals was tested using the Kolmogorov-Smirnov test. If significant departures from normality were observed, the Wilcoxon Rank Sum test was applied. Both ANOVA and ANCOVA were applied to test baseline CSF markers across two or more genotype groups. When ANCOVA was applied, age-at-baseline was included as an additional covariate in the model. If the normality assumption was not satisfied, both ANOVA and ANCOVA models were fitted to the log-transformed data. Box plots were used to visualize data by group.

Relationships between CSF Aβ_42_, CSF p-tau_181_, and other CSF biomarkers, and brain volumes were explored in the overall population and in each genotype group in a simple linear correlation analysis with a Pearson correlation coefficient. The squared Pearson correlation coefficient (*R*^2^) and *P* value were provided in the correlation analysis. Correlation coefficients were defined as: 0.81 ≤ *R*^2^ < 1 as strong; 0.49 ≤ *R*^2^ < 0.81 as moderately strong; 0.25 ≤ *R*^2^ < 0.49 as moderate; 0.09 ≤ *R*^2^ < 0.25 as weak; and R^2^ < 0.09 as negligible. Scatterplots with a simple linear regression line were produced to depict the relationships between two quantitative variables.

Multiple regression analysis was also utilized to assess the functional relationships between biomarkers of interest. This model was applied with CSF Aβ_42_ or CSF p-tau_181_ as the response variable, and *APOEɛ4*, sex, and the biomarker of interest (ie, CSF p-tau_181_ or Aβ_42_, NfL, Ng, YKL-40; hippocampal volume, ventricular volume) as independent variables. This determined the strength of association of CSF Aβ_42_ or CSF p-tau with parameters of interest, in conjunction with the other independent variables included in the model.

## 3. RESULTS

### 3.1 Patient cohort

A total of 102 participants were assessed for eligibility, 56 were excluded, and one enrolled participant did not provide genetic test results. The study sample comprised 45 participants with a mean age of 65.8 years and a mean baseline MMSE total score of 23.6 (**Table S3**). The *APOEɛ4* allele was carried by 33 (73%) participants; 10 (22%) were homozygotes and 23 (51%) were heterozygotes (**Table S3**). All *APOEɛ4* noncarriers were homozygous for *APOEɛ3*, except for one that was *APOEɛ2/ɛ3*. The distribution of *APOE* genotypes (HWE exact *P* value = 1) was consistent with Hardy-Weinberg equilibrium. Participant baseline characteristics were summarized according to *APOEɛ4* genotype and sex (**Tables S3-4**) and have also been described previously (33). The impact of *BCHE-K* allelic status on the phenotype of early AD in *APOEε4* carriers was also assessed and reported (31).

### 3.1 Age at diagnosis and *APOEɛ4* allele status

*APOEɛ4* allele frequency-dependent effects on the risk-of-onset of AD are highest in the seventh decade, wane over 70 years of age, and are particularly reduced after 80 years of age (39–41). Therefore, in this sample of early AD patients aged less than 75 years, the effects of *APOEɛ4* on the AD phenotype may be maximal. In the overall population, the mean age-at-diagnosis of AD was 63.3 years in males and 65.7 years in females. This contrasted with female *APOEɛ4* noncarriers whose mean age-at-diagnosis was 10.7 years later than male *APOEɛ4* noncarriers (**Figure 1A**; 70.4 years in females vs 59.7 years in males, *P* = .009, ANOVA) and nearly 5-fold less time from age-at-diagnosis of AD to study baseline (**Figure 1B**; 4.8 months in females vs 21.6 months in males). The restricted age range for study entry may have been at least partly responsible for the lack of any significant *APOEɛ4* allele frequency-dependent reductions of age-at-diagnosis of AD or age-at-baseline in the overall population, although a trend was observed in females between increasing allele frequency and decreased age (**Figure 1A**).

**Figure 1:**
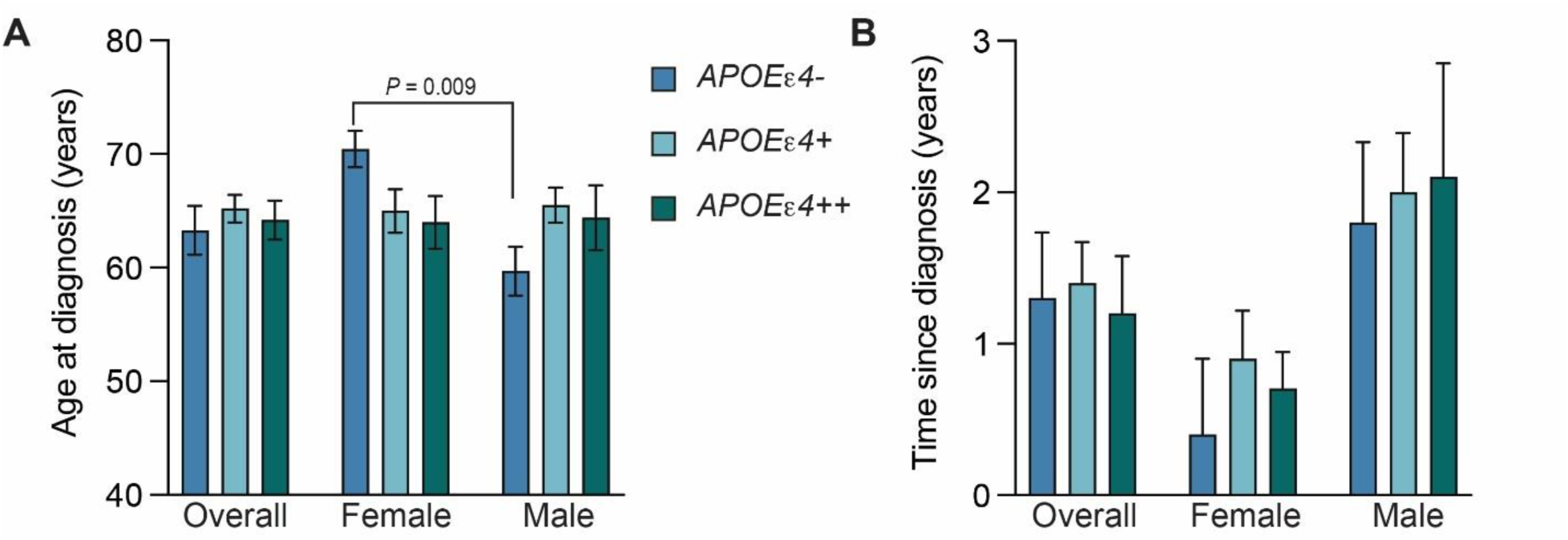
Age at diagnosis and time since diagnosis by *APOEe4* status and sex. The mean (± SEM) was calculated for the age-at-diagnosis (A) and time since diagnosis (B) for all subgroups. Subgroups included the overall population, females, and males, subdivided according to *APOEe4* genotype (blue, *APOEe4* noncarriers; light blue, *APOEe4* heterozygotes; green, *APOEe4* homozygotes). Subgroups and their labels are consistent throughout all figures. Female participants of all genotypes displayed a shorter time since diagnosis than males. See Tables S3 and S4 for more details.

Additionally, female *APOEɛ4* carriers spent a mean of 0.8 years between AD diagnosis and study baseline, whereas male carriers spent 2.0 years (**Figure 1B**), which is consistent with prior studies showing that female *APOEɛ4* carriers less than 75 years of age have a greater risk for AD and a more rapid transition from MCI to AD, especially in heterozygotes (42–44). In *APOEɛ4* homozygotes less than 75 years, the risk for AD may be similar between males and females (45).

### 3.2 *APOEɛ4* allele frequency-dependent changes on CSF markers

*APOEɛ4* allele frequency-dependent increases in amyloid pathology were observed, indexed inversely by CSF Aβ_42_ (**Figure 2A**). The level of amyloid pathology was significantly different across *APOEɛ4* noncarriers, heterozygotes, and homozygotes (**Figure 2A**; CSF Aβ_42_ of 799.2 pg/mL vs 700.2 pg/mL vs 580.7 pg/mL, respectively; *P* = .018 ANOVA, *P* = .024 ANCOVA). The *APOEɛ4* allele frequency-dependent increase in amyloid pathology followed the same trend in males and females but did not reach significance. Male and female *APOEɛ4* homozygotes showed similar levels of amyloid pathology (CSF Aβ_42_ 582.1 vs 579.7 pg/mL, respectively). Higher levels of amyloid pathology are compatible with accumulation beginning at an earlier age in association with *APOEɛ4* allele frequency, with higher levels of amyloid accumulation required in *APOEɛ4/ ɛ4* > *APOEɛ3/ɛ4* > *APOEɛ3/ɛ3* to induce symptoms (46).

**Figure 2.**
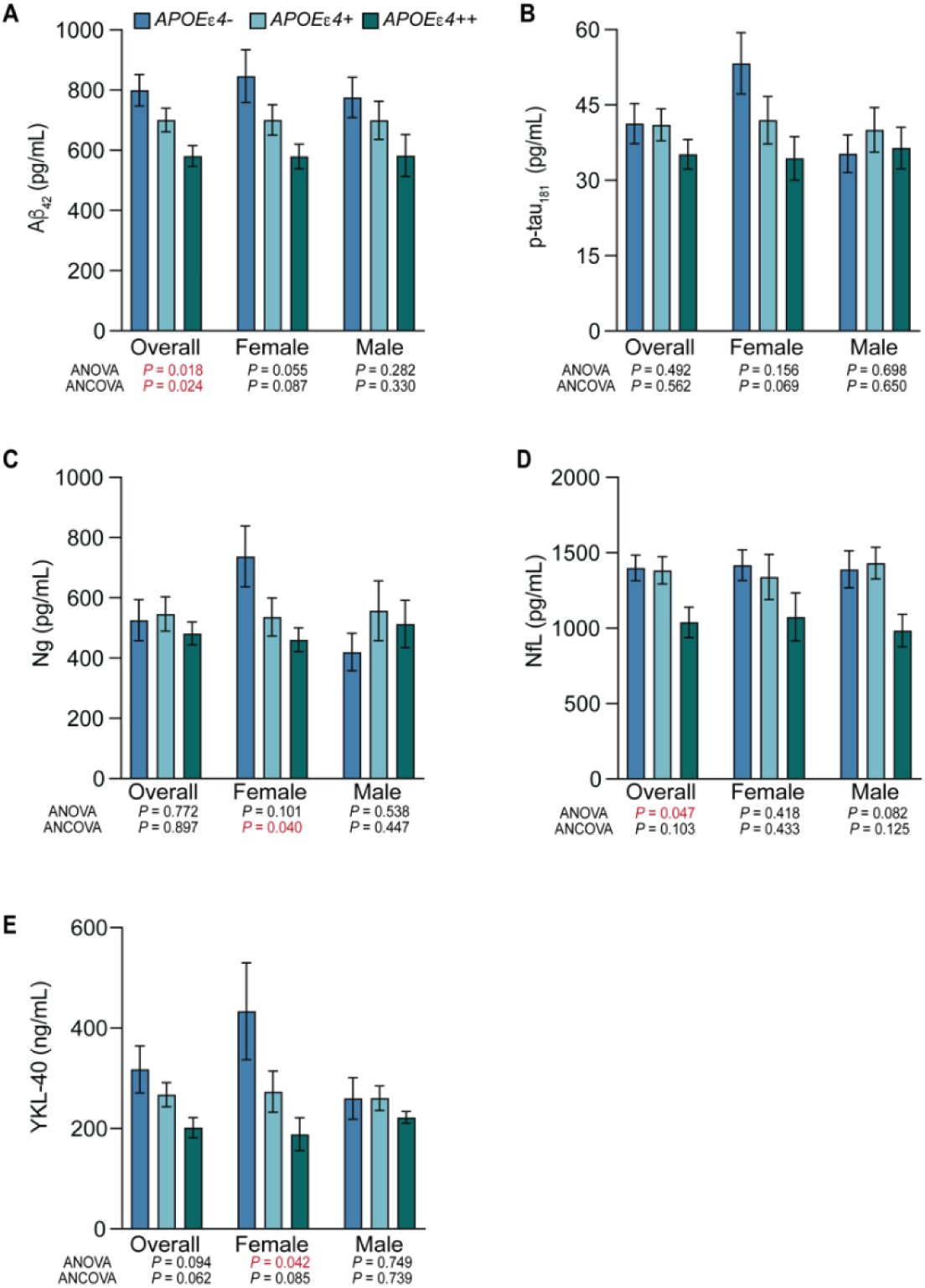
Levels of amyloid pathology, tau pathophysiology, glial activation, and synaptic injury by *APOEε4* allele frequency and sex. The mean and standard error of the mean for CSF Aβ_42_ (A) CSF p-tau_181_ (B) CSF Ng (C) CSF NfL (D) and CSF YKL-40 (E). Females showed significant *APOEe4* allele frequency-dependent relationships for synaptic injury (CSF Ng) and glial activation (CSF YKL-40), and CSF Aβ_42_ and CSF p-tau_181_ showed a trend towards significance. See Tables S3 and S4 for more details.

No significant differences were observed between groups for p-tau_181_. However, *APOEɛ4* homozygotes showed the lowest levels of tau pathology, and in female *APOEɛ4* homozygotes this was most pronounced when compared to female *APOEɛ4* noncarriers (**Figure 2B**; CSF p-tau_181_ of 34.4 pg/mL vs 53.3 pg/mL). Synaptic injury was lowest in *APOEɛ4* homozygotes, and significantly different across female noncarriers, heterozygotes, and homozygotes (**Figure 2C**; CSF Ng of 737.3 pg/mL vs 536.3 pg/mL vs 460.5 pg/mL, respectively; *P* = .101 ANOVA, *P* = .040 ANCOVA). Males showed the opposite trend, where noncarriers exhibited the lowest levels of synaptic injury. Neuroaxonal injury was also significantly different across noncarriers, heterozygotes, and homozygotes (**Figure 2D**; CSF NfL of 1398.9 pg/mL vs 1383.1 pg/mL vs 1038.5 pg/mL, respectively; *P* = .047 ANOVA, *P* = .103 ANCOVA) and followed the same trend in males and females.

Glial activation was highest in noncarriers and lowest in *APOEɛ4* homozygotes (**Figure 2E**). This was especially pronounced in females that showed significant differences across noncarriers, heterozygotes, and homozygotes (CSF YKL-40 of 433.2 ng/mL vs 273.3 ng/mL vs 187.9 ng/mL, respectively; *P* = .042 ANOVA, *P* = .085 ANCOVA).

Overall, *APOEɛ4* homozygotes had the highest levels of amyloid pathology and the lowest levels of tau pathophysiology, neuroaxonal damage, postsynaptic injury, and glial activation (**Figure 2**). This finding was most pronounced in female *APOEɛ4* homozygotes.

### 3.3 Amyloid accumulation, hypofunctional glia mediated clearance, and neurodegenerative pathology

Multiple regression analyses of the overall population identified that amyloid accumulation (inversely indexed by CSF Aβ_42_) was associated with *APOEɛ4* carrier status (*P < .*029), a larger total brain ventricle volume (*P < .*021), less synaptic injury (CSF Ng, *P < .*001), and less tau pathophysiology (CSF p-tau_181_, *P* = .005). There was also a trend towards less glial activation (CSF YKL-40, *P* = .097). In simple linear correlation analyses in the overall population, amyloid pathology showed weak to moderate inverse correlations with tau pathophysiology (*R^2^* = 0.18, *P* = .004), synaptic injury (*R^2^* = 0.26, *P* <.001), and glial activation (*R^2^*= 0.11, *P* = .028) (**Figure 3A-C**). These inverse correlations were absent in *APOEɛ4* noncarriers, but were strengthened in *APOEɛ4* carriers and in males, and in male *APOEɛ4* carriers they were moderate (**Figure 3A-C**). Synaptic injury and amyloid pathology also had a weak inverse correlation in females (*R^2^* = 0.24, *P* = .020), which strengthened to a moderate inverse correlation with in female *APOEɛ4* carriers (*R^2^* = 0.33, *P* = .013) (**Figure 3B**). Amyloid pathology and neuroaxonal damage were not significantly correlated in any subgrouping (**Figure 3D**).

**Figure 3.**
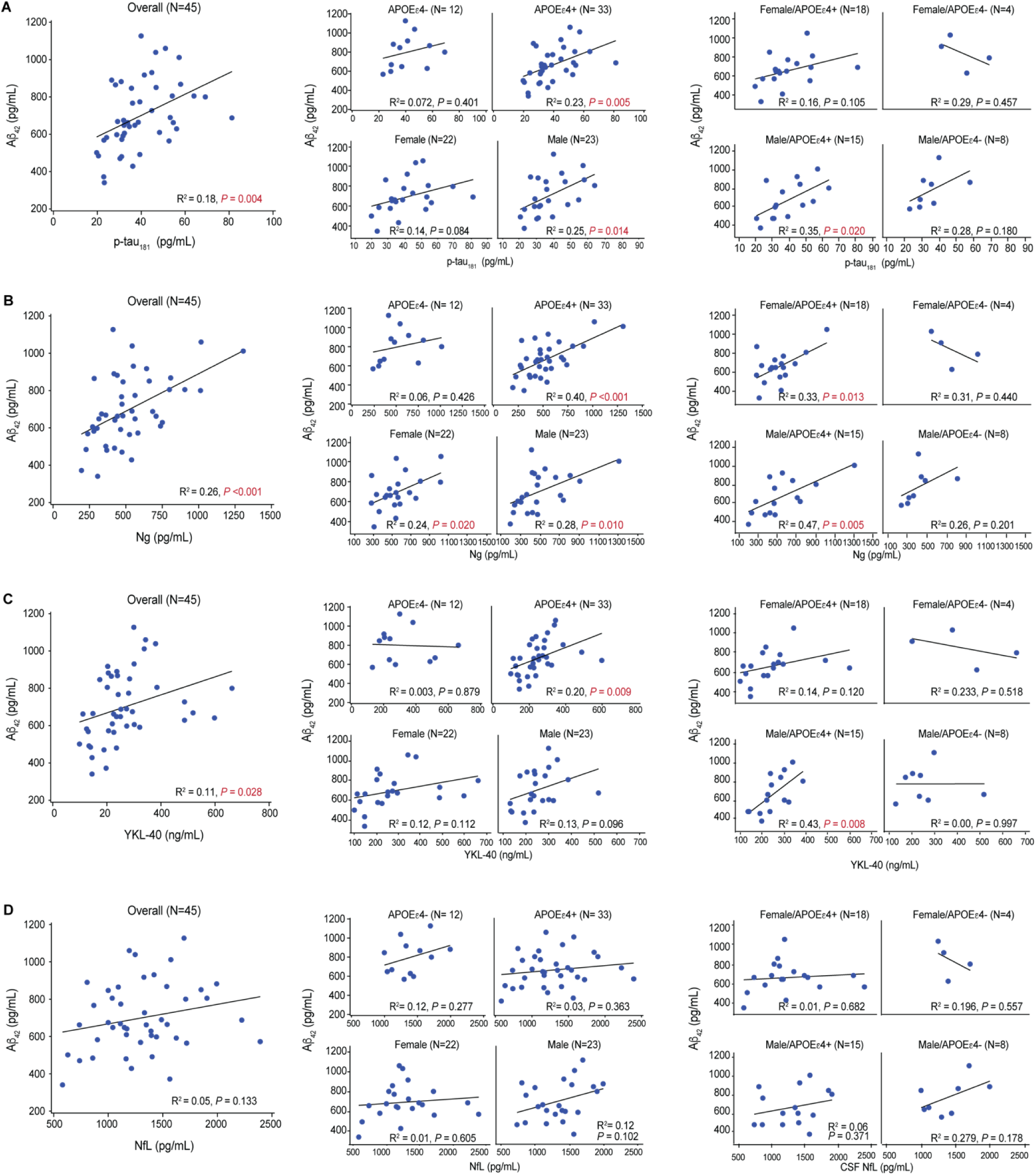
Linear relationships between levels of CSF Aβ_42_ and other CSF markers. Multiple regression identified an association between CSF Aβ_42_ and CSF levels of p-tau_181_, CSF Ng, CSF NfL, and CSF YKL-40. These associations were fit to simple linear regressions to obtain *R*-squared and *P* values. (A) CSF Aβ_42_ (lower levels indicate more amyloid pathology) showed a positive correlation with CSF p-tau_181_ (higher levels indicate more tau pathophysiology) in the overall population, especially in *APOEε4* carriers. When stratified by sex, male *APOEε4* carriers showed the strongest correlation indicating that higher levels of amyloid pathology were associated with less tau pathology. (B) CSF Aβ_42_ showed a positive correlation with Ng (higher levels indicate more synaptic injury) in the overall population, especially in *APOEε4* carriers. When stratified by sex, both male and female *APOEε4* carriers exhibited a strong correlation. (C) CSF Aβ_42_ showed a positive correlation with YKL-40 (higher levels indicate more glial activation) in the overall population, especially in *APOEε4* carriers. When further stratified by sex, male *APOEε4* carriers exhibited the strongest correlation indicating that lower levels of glial activation were associated with more amyloid pathology. (D) CSF Aβ_42_ showed no correlations with NfL (higher levels indicate more axonal injury).

### 3.4 Tau pathophysiology, neurodegenerative pathology, and glial activation

In multiple regression analysis, tau pathophysiology was associated with greater neuroaxonal damage (CSF NfL, *P* = .002), more synaptic injury (CSF Ng, *P < .*001), and higher levels of glial activation (CSF YKL-40, *P* = .01). In simple linear regressions, tau pathophysiology was moderately correlated with neuroaxonal damage in the overall population (*R^2^* = 0.27, *P* < .001) and in *APOEɛ4* carriers (*R^2^* = 0.34, *P* < .001) (**Figure 4A**). In nearly all subgroups, tau pathophysiology showed moderately strong and strong correlations with synaptic injury (**Figure 4B**). Tau pathophysiology was weakly correlated with glial activation in the overall population (*R^2^* = 0.18, *P* = .004) and moderately correlated in *APOEɛ4* noncarriers (*R^2^* = 0.37, *P* = .035) (**Figure 4C**).

**Figure 4.**
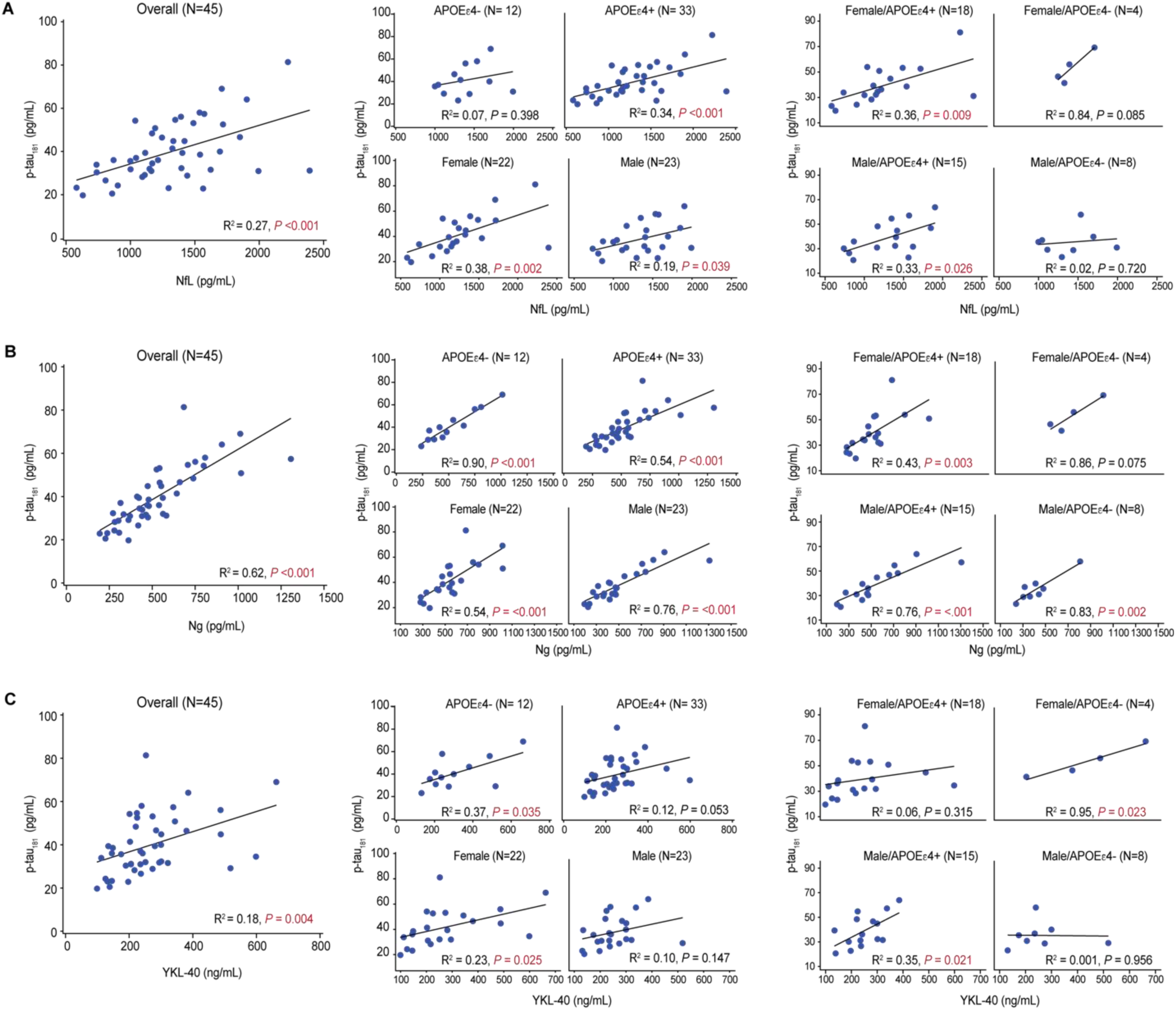
Linear relationships between levels of CSF p-tau_181_ and other CSF markers. Multiple regression identified an association between CSF p-tau_181_ and CSF levels of neurofilament light chain (NfL), Ng, and YKL-40. These associations were fit to simple linear regressions to obtain *R*-squared and *P* values. (A) CSF p-tau_181_ showed a positive correlation with CSF NfL (higher levels indicate more neuroaxonal injury) in the overall population, which was driven by *APOEε4* carriers. When stratified by sex, both male and female *APOEε4* carriers showed a positive correlation. (B) CSF p-tau_181_ showed a positive correlation with Ng (higher levels indicate more synaptic injury) that was moderately strong in the overall population and in *APOEε4* noncarriers and strong in *APOEε4* carriers. This relationship was evident in both males and females, independent of *APOEε4* status. (C) CSF p-tau_181_ showed a positive correlation with YKL-40 (higher levels indicate more glial activation) in the overall population that was stronger in *APOEε4* noncarriers than carriers. When stratified by sex, a positive correlation was present in male *APOEε4* carriers and in female *APOEε4* noncarriers.

Tau pathophysiology and neuroaxonal damage showed a weak correlation in males (*R^2^* = 0.19, *P* = .039) and a moderate correlation in females (*R^2^* = 0.38, *P* = .002) (**Figure 4A**). Tau pathophysiology showed a moderately strong correlation with synaptic injury in both males (*R^2^* = 0.76, *P* = .001) and females (*R^2^* = 0.54, *P* < .001) (**Figure 4B**) and a weak correlation with glial activation in females *(R^2^* = 0.23, *P* = .025) (**Figure 4C**). Notably, correlation of neuroaxonal damage with synaptic injury was significant, but weak in the overall population *(R^2^*= 0.13, *P* = .016), as well as in *APOEɛ4* carriers *(R^2^* = 0.14, *P* = .033) (**Data not shown**). In female *APOEɛ4* noncarriers, correlations between tau and glial activation were significant and strong *(R^2^* = 0.95, *P* = .023), and correlations between tau and neuroaxonal damage or synaptic injury were strong, but did not achieve significance. Whereas, in male *APOEɛ4* noncarriers, correlations of tau with glial activation and neuroaxonal damage were absent (**Figure 4A,C**), and synaptic injury was strong but lacked statistical significance (**Figure 4B**). In female *APOEɛ4* carriers, correlations with tau pathophysiology were absent for glial activation (**Figure 4C**), and moderate for neuroaxonal damage *(R^2^*= 0.36, *P* = .009) and synaptic injury *(R^2^* = 0.43, *P* = .003) (**Figure 4A,B**). In male *APOEɛ4* carriers, tau pathophysiology was moderately correlated with glial activation *(R^2^* = 0.35, *P* = .021) and neuroaxonal damage *(R^2^* = 0.33, *P* = .026) (**Figure 4A,C**), and showed moderately strong correlations with synaptic injury *(R^2^*= 0.76, *P* < .001) (**Figure 4B**).

The most compelling associations of tau pathophysiology with glial activation and neurodegenerative pathology were in female *APOEɛ4* noncarriers and male *APOEɛ4* carriers. Higher CSF p-tau_181_ levels are typically seen in female *APOEɛ4* carriers with preclinical and prodromal AD but not in mild or later dementia stages (9). In *APOEε4* heterozygotes, females may be able to leverage a non-*APOEε4* allele to achieve higher levels of glial activation and increased tau pathology. Whereas increasing levels of tau pathophysiology in male *APOEɛ4* carriers with early AD may reflect more advanced degeneration of the basal forebrain corticolimbic cholinergic neurons, which is indexed by greater limbic-amnestic features and is associated with removal of the cholinergic ‘brake’ on glial function in the MTL followed by wider spread of cortical tau and synaptic pathology. In early AD, spreading cholinergic denervation-induced cortical glial activation-mediated tau and neurodegenerative pathology may be most evident in male *APOEɛ4* homozygotes (32).

### 3.5 Sex and *APOEɛ4* allele influences on cognition

Overall, *APOEɛ4* carriers had a more temporo-limbic (hippocampal atrophy > ventricular expansion) and amnestic (memory > visuospatial impairment) phenotype relative to *APOEɛ4* noncarriers (**Figures 5 and 6**), as described previously (47, 48). In *APOEɛ4* homozygotes, male participants exhibited more of a limbic-amnestic phenotype than females, as evidenced by slightly greater amnestic deficits (**Figure 5B, E**; MMSE memory 4.0 vs 4.2; RBANS delayed memory 48 vs 56; for males vs females) and hippocampal atrophy (**Figure 6A**; 0.22 vs 0.26 %ICV; for males vs females). In *APOEɛ4* heterozygotes, numerically greater amnestic deficits were also observed in male participants (**Figure 5B,E**; MMSE memory 3.8 vs 4.3; RBANS delayed memory 46.9 vs 49.0). These apparent sex differences in *APOEɛ4* heterozygotes were not reflected in amyloid accumulation (**Figure 2A**), tau pathophysiology (**Figure 2B**), hippocampal volumes (**Figure 6A**), or in the mean age-at-diagnosis of AD (65.5 years vs 65.0 years, respectively) (**Figure 1A**).

**Figure 5:**
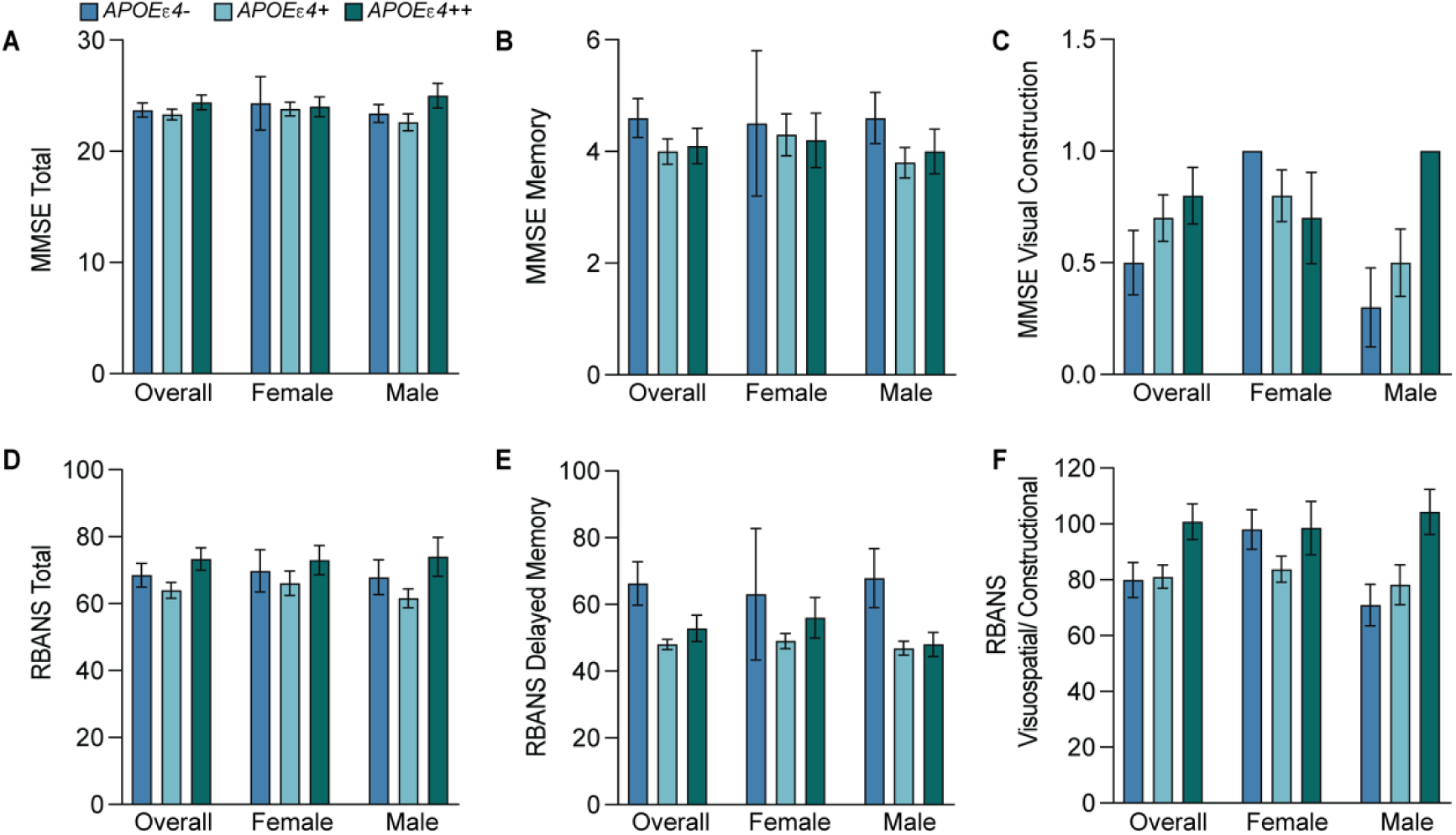
Baseline cognition test scores by *APOEe4* status and sex. The mean and standard error of the mean for baseline cognition tests and two of their individual components: For the MMSE Total (A), Memory (B), and Visual Construction (C); For the RBANS Total (D), Delayed memory (E), and Visuospatial/Constructional (F). Both tests followed the same trends between genotypes with the least overall cognitive and visuospatial deficits in *APOEε4* homozygotes and the greatest amnestic deficits in *APOEε4* carriers. See Tables S3 and S4 for more details.

**Figure 6.**
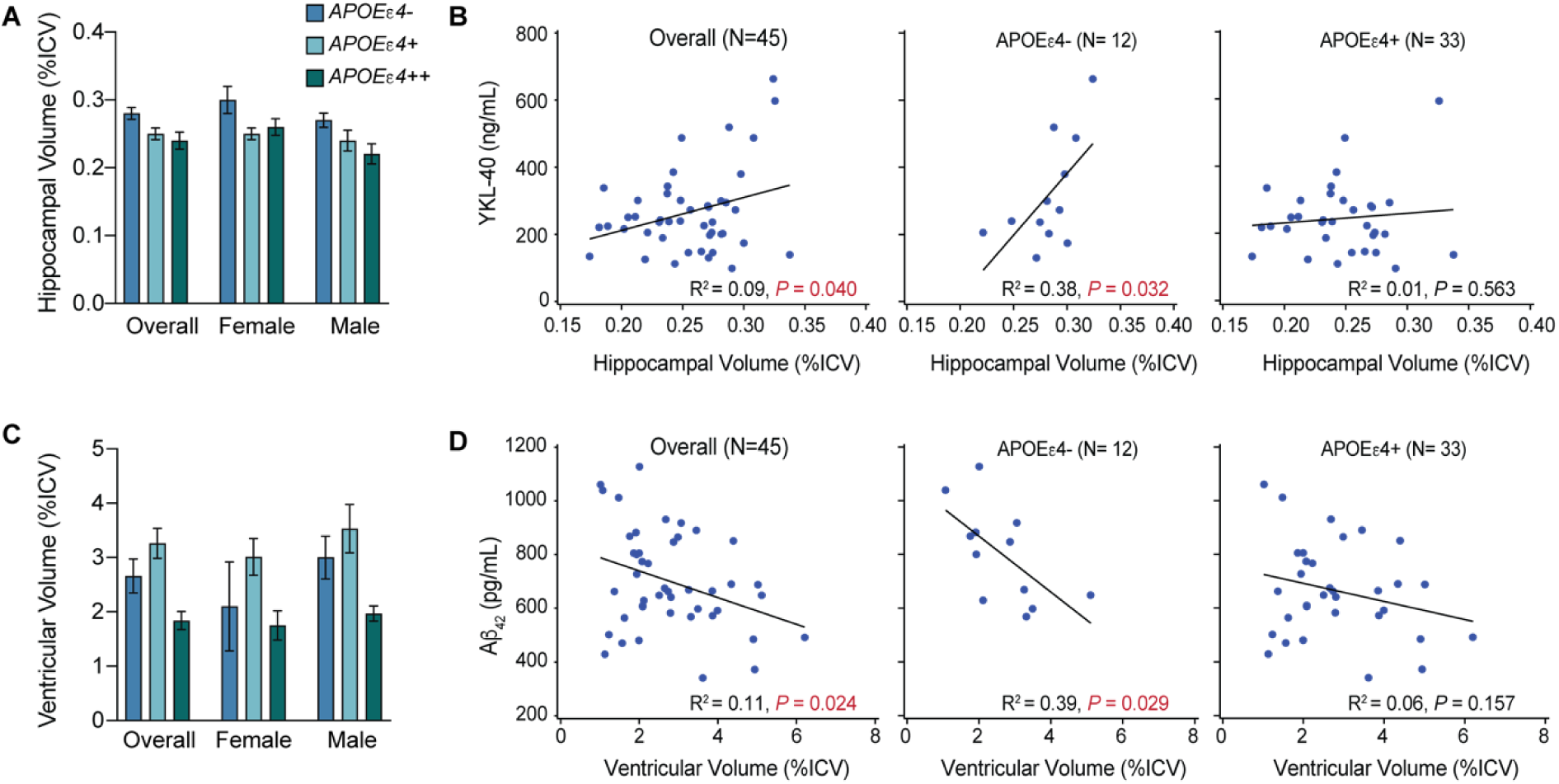
Observations for hippocampal and ventricular volume by *APOEe4* status. The mean and standard error of the mean for hippocampal volume (A) or ventricular volume (C) (as a percentage of total intracranial volume [%ICV]). *R^2^* and *P* values were obtained by fitting simple linear regressions (B,D). CSF YKL-40 correlated with hippocampal volume in the overall population and *APOEε4* noncarriers (B). CSF Aβ_42_ correlated with ventricular volume in the overall population and in *APOEε4* noncarriers (D). See Tables S3 and S4 for more details.

Overall, the limbic-amnestic phenotype was more prominent in male *APOEɛ4* carriers (**Figures 5,6**), suggesting greater and earlier degeneration of the basal forebrain corticolimbic cholinergic projection system in males. Thus, in early-stage AD, male *APOEɛ4* homozygotes may have developed slightly higher levels of glial activation and tau pathophysiology than females because of greater removal of the corticolimbic cholinergic “brake” on glial activation in the MTL and other cortical areas. In male *APOEɛ4* carriers, tau pathophysiology correlated moderately with glial activation *(R^2^*= 0.35, *P* = .021), but this correlation was absent in females (**Figure 4C**). This could indicate that in male *APOEɛ4* carriers with early AD below the age of 75 years, increases in tau pathology are more dependent on limbic and cortical glial activation due to corticolimbic cholinergic denervation. Thus, in early AD, *APOEɛ4* homozygotes may still have the lowest levels of tau pathophysiology, and males and females may be on slightly different journeys to end stage disease where *APOEɛ4* homozygotes will rapidly evolve the greatest burden of tau pathology as they progress through the dementia disease stage continuum (49).

### 3.6 Brain volumes in *APOEɛ4* noncarriers

Neither amyloid nor tau were associated with hippocampal volume. However, total hippocampal volume showed a weak association with glial activation in simple linear correlation analysis (**Figure 6B**): lower glial activation correlated with more hippocampal atrophy. This association was moderate in *APOEɛ4* noncarriers *(R^2^* = 0.38, *P* = .032) and was absent in *APOEɛ4* carriers (**Figure 6B**). This finding aligns with the putative role of glial activation in *APOEɛ4* noncarriers in limiting the accumulation of amyloid pathology and more intact basal forebrain corticolimbic cholinergic projections. Thus, in early AD aged less than 75 years, the absence of apoE4 results in less tau and neurodegenerative pathology in the MTL (48).

Total brain ventricle size showed a weak positive association with amyloid burden in both multiple and simple linear *(R^2^* = 0.11, *P* = .024) correlation analyses (**Figure 6D**). This aligns with weak or equivocal associations between amyloid burden and ventricular expansion or whole brain atrophy in previous studies(50). The association between ventricle size and amyloid burden was moderate in *APOEɛ4* noncarriers *(R^2^* = 0.39, *P* = .029) and was absent in *APOEɛ4* carriers. This suggests that tau and neurodegenerative pathology may be more dependent on levels of amyloid pathology in *APOEɛ4* noncarriers than in carriers. Tau pathophysiology was not correlated with ventricular size.

### 3.7 *APOEɛ4* heterozygotes exhibit influences of both alleles

Relative to homozygotes, *APOEɛ4* heterozygotes had a less extreme limbic-amnestic phenotype, less amyloid, more tau and neurodegenerative pathology, more widespread brain atrophy changes, and higher levels of glial activation (**Figures 2-6**). Relative to noncarriers, heterozygotes also had more amyloid, similar levels of tau and neurodegenerative pathology, and less glial activation. Thus, *APOEɛ4* heterozygotes demonstrate clear influences of both the *APOEɛ4* allele on amyloid accumulation and the non-*APOEɛ4* allele on a more permissive environment for tau spreading. With respect to the limbic-amnestic phenotype, male heterozygotes appear slightly closer to that of all *APOEɛ4* homozygotes, whereas the phenotype of female heterozygotes is slightly closer to all *APOEɛ4* noncarriers.

### 3.8 Hyperfunctional glia in female *APOEɛ4* noncarriers

Post-hoc analysis of *APOEɛ4* noncarriers requires cautious interpretation because of the small number of participants and the preponderance of male (n = 8) relative to female (n = 4) participants. Confirmation is required in prospective analyses of larger populations; however, the subsequent results align well with those of a prodromal AD cohort (32, 44).

Despite the highest burdens of glial activation, tau pathophysiology and synaptic injury (**Figure 2**), female relative to male *APOEɛ4* noncarriers exhibited a later mean age-at-diagnosis of AD of 10.7 years (**Figure 1**) and 5-fold less time from age-at-diagnosis of AD to study baseline. Male *APOEε4* noncarriers had slightly greater amyloid accumulation at baseline but their levels of glial activation, tau pathophysiology, and synaptic injury were markedly lower than in female noncarriers and do not explain their younger age-at-diagnosis of AD. Interestingly, in a 3-4 year study of individuals with prodromal AD, male *APOEɛ4* noncarriers exhibited the slowest decline in brain volumes and the lowest rate of transition to dementia (44). Overall, cognitive impairment in *APOEɛ4* noncarriers was similar in male and female participants, and both had relatively low amnestic deficits. However, males had more dysexecutive clinical features than females, demonstrated by greater deficits in visuospatial/constructional (**Figure 5C,F**; MMSE Visual Construction, 0.3 vs 1.0; RBANS Visuospatial/Constructional, 70.9 vs 98.0) and attentional (MMSE Attention/Calculation, 2.8 and 3.8; RBANS Attention, 71.4 and 81.0, respectively) assessments. Thus, both sexes had relatively non-amnestic cognitive deficits, but female participants exhibited less visuospatial/constructional and attentional impairment, had delayed AD onset (a decade later), and spent less time in the mild AD stage since diagnosis.

In the current study, levels of glial activation and synaptic injury were inversely related to *APOEɛ4* allele frequency in female participants, but this relationship was not seen in male participants (**Figure 2C,E**). The highest levels of glial activation, tau pathophysiology, and synaptic injury were seen in female *APOEɛ4* noncarriers (**Figure 2B,C,E**). Furthermore, correlations of glial activation with tau pathophysiology were weak in the overall population (*R^2^* = 0.18, *P* = .004) and strong in female *APOEɛ4* noncarriers (*R^2^* = 0.95, *P* = .023) (**Figure 4C**). Correlations of synaptic injury with tau pathophysiology were moderately strong in the overall population (*R^2^* = 0.62, *P* < .001) and strong in female *APOEɛ4* noncarriers (*R^2^* = 0.86, *P* = .075) (**Figure 4B**). Correlations of neuroaxonal injury with tau pathophysiology were moderately strong in the overall population (*R^2^* = 0.27, *P* < .001) and strong in female *APOEɛ4* noncarriers (*R^2^*= 0.84, *P* = .085) (**Figure 4A**).

Progression to AD in female *APOEɛ4* noncarriers may only occur after amyloid accumulation eventually reaches the minimum “threshold” to engage the secondary effector tauopathy mechanism when the replication and spread of tau pathology is readily facilitated(51). The substantially greater burden of p-tau_181_ in female *APOEɛ4* noncarriers likely reflects a more diffuse distribution of neocortical tau pathology facilitated by an interaction between sufficient accumulated toxic species of Aβ and a permissive state of glial activation (**Figure 2-4**)(7, 8). In summary, the rapid spread/replication of tau in the cerebral cortex of female *APOEɛ4* noncarriers is strongly associated with higher levels of glial activation, synaptic and neuroaxonal degeneration, and an early AD phenotype of multidomain cognitive impairment with relative preservation of corticolimbic cholinergic innervation, hippocampal volume, and memory function. Female *APOEɛ4* noncarriers in the current study align with a subtype found in 30% of AD patients in a large-scale CSF mass spectrometry proteomic analysis that was characterized by overactive glia (52). This subtype was characterized by increased CSF t-tau and p-tau_181_; relatively widespread cortical atrophy; and increased specific microglial proteins involved in detecting and engulfing amyloid plaque and in innate immune activation, including ApoE and complement complex proteins such as complement component C1q.

Although AD may affect females with more intensity, more rapid brain atrophy, and faster cognitive decline than in males, it does not appear to do so earlier (53, 54). The higher prevalence of AD in females is mostly driven by a divergence of prevalence rates in males and females with increasing age (55, 56). There is an inflection point near 75 years of age where it is difficult to detect significant differences by sex below 75 years, and incidence rates get more divergent above 75 years (57). Most AD below the age of 75 years may be in *APOEɛ4* carriers, with a moderately increased risk in female *APOEɛ4* heterozygotes (45). Interestingly, in *APOE4* carriers, the risk for AD below the age of 75 years is greater in females, but over the age of 75 years the risk for AD is greater in males (39). However, in *APOEɛ4* noncarriers over the age of 75 years, the overall prevalence rates for AD may be substantially higher in females than in males (44). Cortical microglial activation may be higher with age and disproportionately more important for AD progression in females (12). For example, a prior study using neuropathological indicators in human tissue identified that microglial activation in females was associated with both amyloid-mediated increases in tau pathology (∼50%) and direct induction of tau pathology (∼50%), whereas in males, microglial activation was predominantly associated with direct induction of tau pathology (∼74%)(12). In *APOE4* noncarriers, greater CSF tau pathology emerges in females relative to males in prodromal disease and this difference is also present in mild AD (9). In the current study in patients with early AD, tau pathophysiology was substantially greater in female *APOE4* noncarriers (**Figure 2B**).

## 4. DISCUSSION

Genetic variation has a clear influence on the phenotype of early AD in people below the age of 75 years, which was likely made apparent by a decreased likelihood of other age-related pathologies. Amyloid or tau pathology on their own may not pose a substantial risk for neurodegenerative brain pathology, but their co-occurrence may index major risk (58, 59). In *APOEɛ4* homozygotes, deficient glial clearance mechanisms result in early amyloid pathology, however, substantial neurodegeneration and symptoms only begin when tau pathology spreads. In contrast, in female *APOEɛ4* noncarriers, functionally active glia initially limit amyloid pathology, but amyloid accumulation eventually reaches a minimum threshold to trigger tau pathology, resulting in accelerated replication and spread of tau and synaptic pathology. When viewed together, end-stage AD pathology looks similar and may not reflect the divergent pathways taken to get there (49). Through the examination of a younger age group, this study demonstrates that genotype and sex can differentiate phenotypic extremes in early AD.

The amyloid cascade hypothesis of AD implies that parenchymal amyloid positivity at an earlier age should drive secondary effector tau and synaptic pathology, and present with an earlier age-at-onset of AD (51). Although synaptic loss may be necessary to produce clinical impairments, in *APOEɛ4* carriers in the current study, and particularly in males, amyloid pathology was inversely correlated with tau pathophysiology, glial activation and synaptic injury (**Figure 3**). *APOEɛ4* allele frequency-dependent reductions of glial activation were associated with lower Aβ clearance and accumulations of amyloid pathology, but also limited tau pathophysiology and synaptic engulfment (**Figure 7A**). *APOEɛ4* homozygotes had the highest amyloid and the lowest tau and neurodegenerative pathology (**Figure 2**). Tau and neurodegenerative pathology localized in the MTL transition *APOEɛ4* homozygotes from preclinical to clinical AD at an earlier age with less global brain tau and neurodegenerative pathology (**Figure 2**) (48, 60). In younger *APOEɛ4* carriers with global amyloid pathology, tau-PET imaging indicates that tau pathology is more severe with a focal MTL distribution (48, 61). This is associated with more hippocampal atrophy and less global cerebral atrophy.

**Figure 7.**
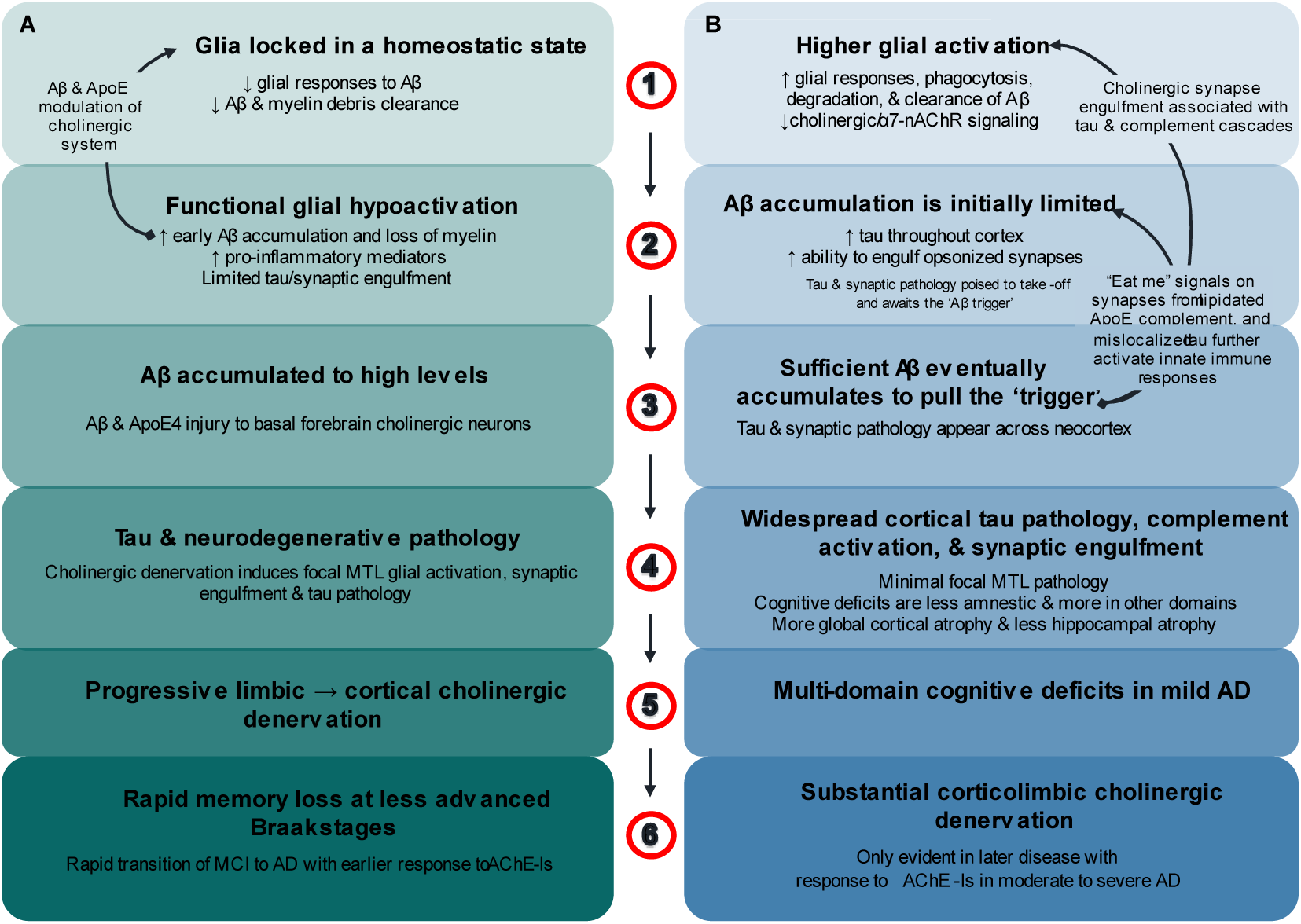
Glial activation differentiates *APOEε4* homozygotes from *APOEε4* noncarriers in early AD. (A) in *APOEε4* homozygotes aged < 75 years, ApoE4 locks glia in a homeostatic state with decreased responsiveness to, and phagocytosis of, extracellular debris including Aβ fibrils (1). “*Functionally underactive*” glia mean earlier and greater amyloid accumulation, but limited synaptic engulfment and loss (2), Aβ accumulates to high levels and along with ApoE4 induces degeneration of basal forebrain corticolimbic cholinergic neurons (3). In prodromal AD, limbic cholinergic denervation removes the cholinergic “brake” on glia to increase glial activation, tau and neurodegenerative pathology in the MTL. This is followed by spread of tau and neurodegenerative pathology beyond the MTL as cholinergic denervation and glial activation progresses to other cortical regions (4) and paralleled by the emergence of a rapidly progressing limbic-amnestic phenotype (5). As corticolimbic cholinergic denervation, glial activation, tau pathology and neurodegeneration spread there is rapid progression from prodromal to dementia stages of AD with an uncharacteristically good response to AChE-Is in mild AD (6). (B) In contrast, female *APOE4* noncarriers usually aged > 75 years have higher levels of glial activation, with induction of microglia response genes that include *APOE* (1). This results in chronically “*functionally over-active*” glia with increased phagocytic and degradative functions that limit Aβ pathology (2). Higher levels of glial activation result in lower levels of amyloid accumulation, but once sufficient Aβ fibrils are present, tau pathology and synaptic engulfment accelerate, and these pathologies spread across the neocortex (3). Generalized cortical tau pathology and synaptic injury index glia tilting through balanced-functionality thresholds to initiate excessive inflammatory and complement system cascades. The lack of focal MTL tau and neurodegenerative pathology means relative sparing of amnestic deficits and hippocampal atrophy, and cortical atrophy changes are more global (4). Multidomain cognitive deficits rapidly transition patients from prodromal to mild stage AD (5). Corticolimbic cholinergic denervation only becomes apparent in the moderate and severe stages of AD where robust AChE-I treatment effects are also observed (6).

Parenchymal amyloid pathology may accumulate to high levels in *APOEɛ4* homozygotes less than 75 years of age before the combined effects of apoE4 and Aβ induce sufficient degeneration of basal forebrain corticolimbic cholinergic neurons to release the ‘brake’ on glial activation. Glial activation, tau, and neurodegenerative pathology first appear in the MTL and then spread to involve other cortical areas as cholinergic denervation becomes more widespread (**Figure 7A**). Across the aging and AD spectrum, *APOEɛ4* carriers present increased microglial activation in early Braak stage regions within the MTL compared to noncarriers, which mediates Aβ-independent effects of *APOEɛ4* on tau accumulation (62). The rapidly progressing limbic-amnestic phenotype indexes the spreading of pathology that is responsible for ushering *APOEɛ4* carriers through prodromal stages of disease and over the dementia threshold, with a response to acetylcholinesterase-inhibitors (AChE-Is) that is apparent in the mild stage of AD (63). Requirements for corticolimbic cholinergic denervation may include apoE4 and low levels of glial activation that induce high levels of accumulated Aβ. However, in most *APOEɛ4* carriers and across the spectrum of AD, an accumulated high amyloid burden inducing cholinergic denervation may not be necessary to induce glial activation if levels of glial activation are already higher for other reasons, such as in *APOEɛ4* heterozygotes and noncarriers, in females, and in patients over the age of about 75 years.

Preclinical *APOEɛ4* carriers exhibit the greatest loss of basal forebrain volume (64). Degeneration of basal forebrain cholinergic neurons that project to the MTL and other cortical structures, precedes and predicts longitudinal MTL degeneration (65). The ascending neuronal projections of the basal forebrain cholinergic system may be particularly vulnerable to the combination of apoE4-mediated glial hypofunction and deficient lipid delivery with high levels of Aβ and tau pathology (66–68). Corticolimbic cholinergic denervation may be evident at early stages of AD (69), and failure of this circuitry is inextricably linked with amnestic deficits (70). Aβ and apoE4 also influence synaptic and extracellular ACh levels (30, 71).

Further lowering of glial activation in *APOEɛ4* carriers, such as in the presence of K variant *BCHE* alleles, induces amyloid accumulation at even younger ages, lowers the age-of-onset of AD, and hastens progression from prodromal to dementia AD, especially in those below the age of 75 years (31, 32, 72, 73). Therefore, factors that increase glial activation, such as aging, may explain why *APOEɛ4* effects on risk for AD may be maximal below the age of 70 years and largely absent above the age of 85 years (39–41). *APOEɛ4* carriers with early AD and a mean age of 70 years show *APOEɛ4* allele frequency-dependent accelerated progression of hippocampal atrophy and decreased global cerebral atrophy, but in individuals more advanced in age or disease progression, brain volume atrophy does not differ by genotype (74–76). In younger *APOEɛ4* homozygotes disseminated neocortical tau pathology and widespread synaptic injury may be initially limited (**Figure 7A**), but as corticolimbic denervation progresses from MTL structures to involve other cortical regions, the spread of tau pathology and synaptic degeneration is facilitated and may be more rapid than in *APOEɛ4* noncarriers. In the current study, tau pathophysiology in *APOEɛ4* carriers had a moderately strong correlation with synaptic injury and moderate correlation with neuroaxonal injury, suggesting that once tau pathology is triggered, it is associated with neurodegeneration (**Figure 4**). In male *APOEɛ4* carriers, moderate to moderately strong correlations of tau pathophysiology with glial activation, synaptic injury, and neuroaxonal damage suggest that increases in glial activation are particularly associated with tau and neurodegenerative pathology in male *APOEɛ4* carriers (**Figure 4**).

In contrast, female *APOEɛ4* noncarriers have high levels of functional glial activation (**Figure 2E**). More effective clearance of Aβ limits amyloid pathology and avoids the substantial accumulation of amyloid at younger ages, however, noncarriers are at increasing risk of developing dementia as they age(32). However, noncarriers are vulnerable because even relatively low levels of amyloid pathology can trigger tau and synaptic pathology to spread rapidly (**Figure 7B**). In the current study, *APOEɛ4* noncarriers with early AD exhibited multi-domain cognitive impairment and global brain atrophy but lacked marked amnestic deficits or hippocampal atrophy. In *APOEɛ4* noncarriers, a significant response to AChE-Is is generally only seen in moderate and severe stages of AD (**Figure 7B**)(32, 63).

*APOEɛ4* allele frequency-dependent reduction of functional glial activation is most evident below the age of 75 years, which contrasts with over-activation of glia in female noncarriers that is most evident greater than 75 years(32). In *APOEɛ4* noncarriers from the current study, tau pathophysiology was strongly correlated with synaptic pathology in both males and females, but correlations with glial activation and neuroaxonal damage were only strong in females and absent in males (**Figure 4A-C**). In addition, females, but not males, showed an inverse *APOEɛ4* allele frequency-dependent relationships with glial activation, tau pathophysiology and synaptic injury (**Figure 2**). The highest levels of glial activation, tau and synaptic pathology, and the lowest levels of amyloid pathology were evidenced in female *APOEɛ4* noncarriers (**Figure 2**). In *APOEɛ4* noncarriers, sex had a major effect on age-of-diagnosis of AD, with a significantly earlier mean age in male relative to female participants of 10.7 years. Thus, while female *APOEɛ4* noncarriers may be relatively resilient at younger ages, their risk for AD may increase with age becoming particularly divergent from males over the age of 75 years (57). This is supported by the literature, where 40% of female *APOEɛ4* noncarriers aged 75 years or more with prodromal AD, transitioned to dementia during a four-year study compared to only 21% of male noncarriers (44).

Limitations of this current study include its retrospective nature, its small size, and that participants were largely of European ancestry. In addition, discerning clinical phenotypes on a variable background of AChE-I therapy (**Tables S3, 4**) may be problematic because *APOEɛ4* carriers are more responsive to AChE-I treatment in the mild stage of AD, therefore attention, processing speed, and amnestic deficits may have been partly obscured (63).

In early AD below 75 years of age, increased *APOEɛ4* allele frequency is associated with decreased functional activation of glia; this was evident in females, but not in males. Male and female *APOEε4* homozygotes had the lowest levels of glial activation, while female *APOEɛ4* noncarriers, and to a lesser extent female *APOEɛ4* heterozygotes, had increased levels of glial activation. These age, genotype, and sex influences on levels of glial activation are important determinants in the mix and timing of amyloid and tau pathology, neurodegeneration, denervation of the corticolimbic cholinergic system, and clinical features of early AD. They may explain much of the phenotypic heterogeneity in early AD populations aged below 75 years. These findings should be confirmed in larger, prospective, and longitudinal studies.

## Data Availability

All data produced in the present study are available upon reasonable request to the authors

## Acknowledgements

We thank the participants and their companions who participated in the study; the sites, and study team from Ionis for executing the study; and Gwendolyn Kaeser who created the figures, and edited and styled the manuscript per journal requirements.

## Author contributions

RML was responsible for study design, statistical analysis plan design, and data interpretation, and wrote the manuscript. DL advised on statistical analysis plan, performed data analysis, data interpretation, and performed critical review of the manuscript. TDS advised on structure of the manuscript, data interpretation, and performed critical review of the manuscript.

## Availability of data and materials

The baseline data that support the findings of this investigation are available in the supplementary materials (Tables S3–S4) and additional data and materials are available upon request.

## Declarations

Ethics approval and consent to participate

Patients provided written, informed consent at the time of recruitment. The study was approved by the institutional review board or independent ethics committee at each investigational site.

## References

1. Yamazaki Y, Zhao N, Caulfield TR, Liu C-C, Bu G. Apolipoprotein E and Alzheimer disease: pathobiology and targeting strategies. Nature Reviews Neurology. 2019;15(9):501–18.

2. Koutsodendris N, Nelson MR, Rao A, Huang Y. Apolipoprotein E and Alzheimer’s Disease: Findings, Hypotheses, and Potential Mechanisms. Annu Rev Pathol. 2022;17:73–99.

3. Guillot-Sestier MV, Doty KR, Gate D, Rodriguez J, Jr., Leung BP, Rezai-Zadeh K, Town T. Il10 deficiency rebalances innate immunity to mitigate Alzheimer-like pathology. Neuron. 2015;85(3):534–48.

4. Suarez-Calvet M, Morenas-Rodriguez E, Kleinberger G, Schlepckow K, Araque Caballero MA, Franzmeier N, et al. Early increase of CSF sTREM2 in Alzheimer’s disease is associated with tau related-neurodegeneration but not with amyloid-beta pathology. Mol Neurodegener. 2019;14(1):1.

5. Jain N, Lewis CA, Ulrich JD, Holtzman DM. Chronic TREM2 activation exacerbates Abeta-associated tau seeding and spreading. J Exp Med. 2023;220(1).

6. Gratuze M, Leyns CE, Sauerbeck AD, St-Pierre MK, Xiong M, Kim N, et al. Impact of TREM2R47H variant on tau pathology-induced gliosis and neurodegeneration. J Clin Invest. 2020;130(9):4954–68.

7. Pascoal TA, Benedet AL, Ashton NJ, Kang MS, Therriault J, Chamoun M, et al. Microglial activation and tau propagate jointly across Braak stages. Nature medicine. 2021;27(9):1592–9.

8. Bellaver B, Povala G, Ferreira PCL, Ferrari-Souza JP, Leffa DT, Lussier FZ, et al. Astrocyte reactivity influences amyloid-β effects on tau pathology in preclinical Alzheimer’s disease. Nature medicine. 2023.

9. Mofrad RB, Tijms BM, Scheltens P, Barkhof F, van der Flier WM, Sikkes SA, Teunissen CEJN. Sex differences in CSF biomarkers vary by Alzheimer disease stage and APOE ε4 genotype. 2020;95(17):e2378–e88.

10. Sala Frigerio C, Wolfs L, Fattorelli N, Thrupp N, Voytyuk I, Schmidt I, et al. The Major Risk Factors for Alzheimer’s Disease: Age, Sex, and Genes Modulate the Microglia Response to Aβ Plaques. Cell Rep. 2019;27(4):1293–306.e6.

11. Berchtold NC, Cribbs DH, Coleman PD, Rogers J, Head E, Kim R, et al. Gene expression changes in the course of normal brain aging are sexually dimorphic. Proc Natl Acad Sci U S A. 2008;105(40):15605–10.

12. Casaletto KB, Nichols E, Aslanyan V, Simone SM, Rabin JS, La Joie R, et al. Sex-specific effects of microglial activation on Alzheimer’s disease proteinopathy in older adults. Brain. 2022;145(10):3536–45.

13. Yin Z, Rosenzweig N, Kleemann KL, Zhang X, Brandão W, Margeta MA, et al. APOE4 impairs the microglial response in Alzheimer’s disease by inducing TGFβ-mediated checkpoints. Nat Immunol. 2023;24(11):1839–53.

14. Machlovi SI, Neuner SM, Hemmer BM, Khan R, Liu Y, Huang M, et al. APOE4 confers transcriptomic and functional alterations to primary mouse microglia. Neurobiology of disease. 2022;164:105615.

15. Podleśny-Drabiniok A, Marcora E, Goate AM. Microglial Phagocytosis: A Disease-Associated Process Emerging from Alzheimer’s Disease Genetics. Trends Neurosci. 2020;43(12):965–79.

16. Safaiyan S, Kannaiyan N, Snaidero N, Brioschi S, Biber K, Yona S, et al. Age-related myelin degradation burdens the clearance function of microglia during aging. Nat Neurosci. 2016;19(8):995–8.

17. Lin YT, Seo J, Gao F, Feldman HM, Wen HL, Penney J, et al. APOE4 Causes Widespread Molecular and Cellular Alterations Associated with Alzheimer’s Disease Phenotypes in Human iPSC-Derived Brain Cell Types. Neuron. 2018;98(6):1141–54.e7.

18. Hu Y, Fryatt GL, Ghorbani M, Obst J, Menassa DA, Martin-Estebane M, et al. Replicative senescence dictates the emergence of disease-associated microglia and contributes to Aβ pathology. Cell Rep. 2021;35(10):109228.

19. Connolly K, Lehoux M, O’Rourke R, Assetta B, Erdemir GA, Elias JA, et al. Potential role of chitinase-3-like protein 1 (CHI3L1/YKL-40) in neurodegeneration and Alzheimer’s disease. Alzheimers Dement. 2022.

20. Yin F. Lipid metabolism and Alzheimer’s disease: clinical evidence, mechanistic link and therapeutic promise. Febs j. 2023;290(6):1420–53.

21. Leng L, Yuan Z, Pan R, Su X, Wang H, Xue J, et al. Microglial hexokinase 2 deficiency increases ATP generation through lipid metabolism leading to β-amyloid clearance. Nature metabolism. 2022;4(10):1287–305.

22. Choi H, Mook-Jung I. Lipid fuel for hungry-angry microglia. Nature metabolism. 2022;4(10):1223–4.

23. Prakash P, Manchanda P, Paouri E, Bisht K, Sharma K, Wijewardhane PR, et al. Amyloid β Induces Lipid Droplet-Mediated Microglial Dysfunction in Alzheimer’s Disease. bioRxiv. 2023:2023.06.04.543525.

24. Baenziger JE, Morris ML, Darsaut TE, Ryan SE. Effect of membrane lipid composition on the conformational equilibria of the nicotinic acetylcholine receptor. J Biol Chem. 2000;275(2):777–84.

25. Barrantes FJ. Cholesterol effects on nicotinic acetylcholine receptor: cellular aspects. Subcell Biochem. 2010;51:467–87.

26. Benfante R, Di Lascio S, Cardani S, Fornasari D. Acetylcholinesterase inhibitors targeting the cholinergic anti-inflammatory pathway: a new therapeutic perspective in aging-related disorders. Aging Clin Exp Res. 2021;33(4):823–34.

27. Kumar R, Nordberg A, Darreh-Shori T. Amyloid-β peptides act as allosteric modulators of cholinergic signalling through formation of soluble BAβACs. Brain. 2016;139(Pt 1):174–92.

28. Baidya AT, Kumar A, Kumar R, Darreh-Shori T. Allosteric Binding Sites of Aβ Peptides on the Acetylcholine Synthesizing Enzyme ChAT as Deduced by In Silico Molecular Modeling. Int J Mol Sci. 2022;23(11).

29. Darreh-Shori T, Siawesh M, Mousavi M, Andreasen N, Nordberg A. Apolipoprotein epsilon4 modulates phenotype of butyrylcholinesterase in CSF of patients with Alzheimer’s disease. Journal of Alzheimer’s disease : JAD. 2012;28(2):443–58.

30. Darreh-Shori T, Vijayaraghavan S, Aeinehband S, Piehl F, Lindblom RP, Nilsson B, et al. Functional variability in butyrylcholinesterase activity regulates intrathecal cytokine and astroglial biomarker profiles in patients with Alzheimer’s disease. Neurobiology of aging. 2013;34(11):2465–81.

31. Lane RM, Darreh-Shori T, Junge C, Li D, Yang QM, Edwards A, et al. Onset of Alzheimer disease in apolipoprotein ɛ4 carriers is earlier in butyrylcholinesterase K variant carriers. medRxiv. 2024.

32. Lane RM, Darreh-Shori T. Understanding the beneficial and detrimental effects of donepezil and rivastigmine to improve their therapeutic value. Journal of Alzheimer’s disease : JAD. 2015;44(4):1039–62.

33. Mummery CJ, Borjesson-Hanson A, Blackburn DJ, Vijverberg EGB, De Deyn PP, Ducharme S, et al. Tau-targeting antisense oligonucleotide MAPT(Rx) in mild Alzheimer’s disease: a phase 1b, randomized, placebo-controlled trial. Nature medicine. 2023.

34. Folstein MF, Folstein SE, McHugh PR. "Mini-mental state". A practical method for grading the cognitive state of patients for the clinician. J Psychiatr Res. 1975;12(3):189–98.

35. Morris JC, Ernesto C, Schafer K, Coats M, Leon S, Sano M, et al. Clinical dementia rating training and reliability in multicenter studies: the Alzheimer’s Disease Cooperative Study experience. Neurology. 1997;48(6):1508–10.

36. Albert MS, DeKosky ST, Dickson D, Dubois B, Feldman HH, Fox NC, et al. The diagnosis of mild cognitive impairment due to Alzheimer’s disease: recommendations from the National Institute on Aging-Alzheimer’s Association workgroups on diagnostic guidelines for Alzheimer’s disease. Alzheimers Dement. 2011;7(3):270–9.

37. Wang X, Ghayoor A, Novicki A, Holmes S, Seibyl J, Hesterman J. [P4–266]: APPLICATION OF A MULTI-ATLAS SEGMENTATION TOOL TO HIPPOCAMPUS, VENTRICLE AND WHOLE BRAIN SEGMENTATION. Alzheimer’s & Dementia. 2017;13(7S_Part_28):P1385–P6.

38. Graffelman J. Exploring diallelic genetic markers: the HardyWeinberg package. Journal of Statistical Software. 2015;64:1–23.

39. Huang LC, Lee MY, Chien CF, Chang YP, Li KY, Yang YH. Age and sex differences in the association between APOE genotype and Alzheimer’s disease in a Taiwan Chinese population. Frontiers in aging neuroscience. 2023;15:1246592.

40. Bellou E, Baker E, Leonenko G, Bracher-Smith M, Daunt P, Menzies G, et al. Age-dependent effect of APOE and polygenic component on Alzheimer’s disease. Neurobiology of aging. 2020;93:69–77.

41. Whitwell JL, Tosakulwong N, Weigand SD, Graff-Radford J, Ertekin-Taner N, Machulda MM, et al. Relationship of APOE, age at onset, amyloid and clinical phenotype in Alzheimer disease. 2021;108:90–8.

42. Wang X, Zhou W, Ye T, Lin X, Zhang J. Sex Difference in the Association of APOE4 with Memory Decline in Mild Cognitive Impairment. Journal of Alzheimer’s disease : JAD. 2019;69(4):1161–9.

43. Neu SC, Pa J, Kukull W, Beekly D, Kuzma A, Gangadharan P, et al. Apolipoprotein E Genotype and Sex Risk Factors for Alzheimer Disease: A Meta-analysis. JAMA neurology. 2017;74(10):1178–89.

44. Lane RM, He Y. Butyrylcholinesterase genotype and gender influence Alzheimer’s disease phenotype. Alzheimers Dement. 2013;9(2):e1–73.

45. Farrer LA, Cupples LA, Haines JL, Hyman B, Kukull WA, Mayeux R, et al. Effects of Age, Sex, and Ethnicity on the Association Between Apolipoprotein E Genotype and Alzheimer Disease: A Meta-analysis. JAMA. 1997;278(16):1349–56.

46. Fleisher AS, Chen K, Liu X, Ayutyanont N, Roontiva A, Thiyyagura P, et al. Apolipoprotein E ε4 and age effects on florbetapir positron emission tomography in healthy aging and Alzheimer disease. Neurobiology of aging. 2013;34(1):1–12.

47. Groot C, Grothe MJ, Mukherjee S, Jelistratova I, Jansen I, van Loenhoud AC, et al. Differential patterns of gray matter volumes and associated gene expression profiles in cognitively-defined Alzheimer’s disease subgroups. NeuroImage Clinical. 2021;30:102660.

48. Mattsson N, Ossenkoppele R, Smith R, Strandberg O, Ohlsson T, Jögi J, et al. Greater tau load and reduced cortical thickness in APOE ε4-negative Alzheimer’s disease: a cohort study. Alzheimers Res Ther. 2018;10(1):77.

49. Reiman EM, Arboleda-Velasquez JF, Quiroz YT, Huentelman MJ, Beach TG, Caselli RJ, et al. Exceptionally low likelihood of Alzheimer’s dementia in APOE2 homozygotes from a 5,000-person neuropathological study. Nat Commun. 2020;11(1):667.

50. Josephs KA, Whitwell JL, Ahmed Z, Shiung MM, Weigand SD, Knopman DS, et al. Beta-amyloid burden is not associated with rates of brain atrophy. Ann Neurol. 2008;63(2):204–12.

51. Hardy JA, Higgins GA. Alzheimer’s disease: the amyloid cascade hypothesis. Science. 1992;256(5054):184-5.

52. Tijms BM, Vromen EM, Mjaavatten O, Holstege H, Reus LM, Lee Svd, et al. Large-scale cerebrospinal fluid proteomic analysis in Alzheimer’s disease patients reveals five molecular subtypes with distinct genetic risk profiles. medRxiv. 2023:2023.05.10.23289793.

53. Sauty B, Durrleman S. Impact of sex and APOE-ε4 genotype on patterns of regional brain atrophy in Alzheimer’s disease and healthy aging. Front Neurol. 2023;14:1161527.

54. Laws KR, Irvine K, Gale TM. Sex differences in cognitive impairment in Alzheimer’s disease. World J Psychiatry. 2016;6(1):54–65.

55. Ferretti MT, Iulita MF, Cavedo E, Chiesa PA, Schumacher Dimech A, Santuccione Chadha A, et al. Sex differences in Alzheimer disease - the gateway to precision medicine. Nat Rev Neurol. 2018;14(8):457–69.

56. Dubal DB. Sex difference in Alzheimer’s disease: An updated, balanced and emerging perspective on differing vulnerabilities. Handb Clin Neurol. 2020;175:261–73.

57. Beam CR, Kaneshiro C, Jang JY, Reynolds CA, Pedersen NL, Gatz M. Differences Between Women and Men in Incidence Rates of Dementia and Alzheimer’s Disease. Journal of Alzheimer’s disease : JAD. 2018;64(4):1077–83.

58. Rhein V, Song X, Wiesner A, Ittner LM, Baysang G, Meier F, et al. Amyloid-beta and tau synergistically impair the oxidative phosphorylation system in triple transgenic Alzheimer’s disease mice. Proc Natl Acad Sci U S A. 2009;106(47):20057–62.

59. Puzzo D, Argyrousi EK, Staniszewski A, Zhang H, Calcagno E, Zuccarello E, et al. Tau is not necessary for amyloid-β–induced synaptic and memory impairments. The Journal of Clinical Investigation. 2020;130(9):4831–44.

60. Ossenkoppele R, Binette AP, Groot C, Smith R, Strandberg O, Palmqvist S, et al. Amyloid and Tau PET positive cognitively unimpaired individuals: Destined to decline? 2022:2022.05.23.22275241.

61. Susanto TA, Pua EP, Zhou J. Cognition, brain atrophy, and cerebrospinal fluid biomarkers changes from preclinical to dementia stage of Alzheimer’s disease and the influence of apolipoprotein e. Journal of Alzheimer’s disease : JAD. 2015;45(1):253–68.

62. Ferrari-Souza JP, Lussier FZ, Leffa DT, Therriault J, Tissot C, Bellaver B, et al. APOEε4 associates with microglial activation independently of Aβ plaques and tau tangles. Sci Adv. 2023;9(14):eade1474.

63. Lane RM, He Y. Emerging hypotheses regarding the influences of butyrylcholinesterase-K variant, APOE epsilon 4, and hyperhomocysteinemia in neurodegenerative dementias. Medical hypotheses. 2009;73(2):230–50.

64. Schmitz TW, Soreq H, Poirier J, Spreng RN. Longitudinal Basal Forebrain Degeneration Interacts with TREM2/C3 Biomarkers of Inflammation in Presymptomatic Alzheimer’s Disease. J Neurosci. 2020;40(9):1931–42.

65. Schmitz TW, Nathan Spreng R. Basal forebrain degeneration precedes and predicts the cortical spread of Alzheimer’s pathology. Nat Commun. 2016;7:13249.

66. Yu MC, Chuang YF, Wu SC, Ho CF, Liu YC, Chou CJ. White matter hyperintensities in cholinergic pathways are associated with dementia severity in e4 carriers but not in non-carriers. Front Neurol. 2023;14:1100322.

67. Hu L, Wong TP, Cote SL, Bell KF, Cuello AC. The impact of Abeta-plaques on cortical cholinergic and non-cholinergic presynaptic boutons in alzheimer’s disease-like transgenic mice. Neuroscience. 2003;121(2):421–32.

68. Bell KF, Claudio Cuello A. Altered synaptic function in Alzheimer’s disease. European journal of pharmacology. 2006;545(1):11–21.

69. Mesulam M, Shaw P, Mash D, Weintraub S. Cholinergic nucleus basalis tauopathy emerges early in the aging-MCI-AD continuum. Ann Neurol. 2004;55(6):815–28.

70. Ballinger EC, Ananth M, Talmage DA, Role LW. Basal Forebrain Cholinergic Circuits and Signaling in Cognition and Cognitive Decline. Neuron. 2016;91(6):1199–218.

71. Fontana IC, Kumar A, Nordberg A. The role of astrocytic alpha7 nicotinic acetylcholine receptors in Alzheimer disease. Nat Rev Neurol. 2023;19(5):278–88.

72. Chuang YF, Varma V, An Y, Tanaka T, Davatzikos C, Resnick SM, Thambisetty M. Interaction between Apolipoprotein E and Butyrylcholinesterase Genes on Risk of Alzheimer’s Disease in a Prospective Cohort Study. Journal of Alzheimer’s disease : JAD. 2020;75(2):417–27.

73. Lane R, Feldman HH, Meyer J, He Y, Ferris SH, Nordberg A, et al. Synergistic effect of apolipoprotein E epsilon4 and butyrylcholinesterase K-variant on progression from mild cognitive impairment to Alzheimer’s disease. Pharmacogenetics and genomics. 2008;18(4):289–98.

74. Bigler ED, Lowry CM, Anderson CV, Johnson SC, Terry J, Steed MJAJoN. Dementia, quantitative neuroimaging, and apolipoprotein E genotype. 2000;21(10):1857–68.

75. Yasuda M, Mori E, Kitagaki H, Yamashita H, Hirono N, Shimada K, et al. Apolipoprotein E ε4 allele and whole brain atrophy in late-onset Alzheimer’s disease. 1998;155(6):779–84.

76. Jack Jr CR, Petersen RC, Xu YC, O’Brien PC, Waring SC, Tangalos EG, et al. Hippocampal atrophy and apolipoprotein E genotype are independently associated with Alzheimer’s disease. 1998;43(3):303–10.

