## Supplementary tables for "Glial activation mediates phenotypic effects of *APOEε4* and sex in Alzheimer’s disease"

### Supplementary Materials

#### Supplemental Tables

**Table S1.** Ethics committees approving clinical study.

**Table S2.** Description of CSF biomarkers of AD pathology, glial activation, neuroaxonal damage, and synaptic injury.

**Table S3.** Mild AD phenotype across genotype groups defined by *APOE4* allele frequency.

**Table S4.** Mild AD phenotype in *APOE4* carriers by *APOE4* allele frequency and sex.

### Supplemental Tables

Table S1. Ethics committees approving clinical study.

Ethics committees approving clinical study

| Principal Investigator | IRB/Ethic Committee | Reference number |
|---|---|---|
| Dr Albert Ludolph | Ethikkommission der Universität Ulm | 412/16 |
| Dr Siegfried Muhlack | Ethik-Kommission der Ruhr-Universität Bochum | 16-5929 |
| Dr Catherine Mummery | London-Central Research Ethics Committee Manchester HRA Centre | 17/LO/0440 |
| Dr Simon Ducharme | MUHC Neurosciences Research Ethics Board | 2017-3206 |
| Dr Juha Rinne | National Committee on Medical Research Ethics | 73/06.00.01/2017 |
| Dr Ralf Bodenschatz | Ethikkommission der Sächsischen Landesärztekammer | EK-AMG-MCB-155/16-1 |
| Dr Peter Paul de Deyn | Central Committee on Research Involving Human Subjects | NL60032.000.16 |
| Dr Anne Borjesson Hansen | Regionala etikprövningsnämnden i Stockholm Karolinska Institutet i Solna | 2017/300-31 |
| Dr Michael Jonsson | Regionala etikprövningsnämnden i Stockholm Karolinska Institutet i Solna | 2017/300-31 |
| Dr Daniel Blackburn | London-Central Research Ethics Committee Manchester HRA Centre | 17/LO/0440 |
| Dr Anja Schneider | Ethikkommission an der Medizinischen Fakultät der Rheinischen Friedrich-Wilhelms-Universität Bonn | 035/18-AMG |
| Dr Phillipus Scheltens | Central Committee on Research Involving Human Subjects | NL60032.000.16 |

**Table S2. Description of CSF biomarkers of AD pathology, glial activation, neuroaxonal damage, and synaptic injury.**

| CSF analyte | Biology indexed |
|---|---|
| **A$\beta_{42}$ (**β-amyloid 1-42)<br>Elecsys β-amyloid 1-42 CSF performed at Roche Diagnostics, Indianapolis, IN | Parenchymal amyloid accumulation |
| **p-tau$_{181}$** (tau phosphorylated at threonine 181)<br>Elecsys Phospho-Tau (181P) CSF performed at Roche Diagnostics, Indianapolis, IN | Tau pathophysiology. CSF p-tau$_{181}$ may reflect a mix of amyloid and tau pathological changes in the brain (75), and is therefore not a "pure" marker of tau tangle load in the brain. |
| **NfL*** (neurofilament light chain)<br>Uman, performed at Immunologix, Tampa, FL | Neuroaxonal degeneration |
| **Ng***(neurogranin)<br>Euroimmune, performed at Immunologix, Tampa, FL) | Postsynaptic injury |
| **YKL-40*** (chitinase-3-like protein 1)<br>ELLA (Protein Simple), performed at Immunologix, Tampa, FL) | Putative master regulator of microglial and astrocyte activation (19). YKL-40 is a context-dependent modulator of glial phagocytic activity in both mice and humans. The associations of this biomarker with transcriptional, morphological, and functional states of glia are not clear and clarity may require large multiomic datasets and machine learning. |

*All CSF samples for NfL, Ng, and YKL-40 were tested using a single batch of reagents.

#### Table S3. Mild AD phenotype across *APOE4* allele frequency groups.

| Variable | | | 2 *APOEε4* (N = 10) | 1 *APOEε4* (N = 23) | No *APOEε4* (N = 12) | 1 or 2 *APOEε4* (N = 33) | Overall (N=45) |
|---|---|---|---|---|---|---|---|
| **Sex** | Female | N (%) | 6 (60.0%) | 12 (52.2%) | 4 (33.3%) | 18 (54.5%) | 22 (48.9%) |
| | Male | N (%) | 4 (40.0%) | 11 (47.8%) | 8 (66.7%) | 15 (45.5%) | 23 (51.1%) |
| **Onset of AD** | Age-at-diagnosis (yrs) | **Mean (SD, SEM)** | **64.2 (5.4, 1.7)** | **65.2 (5.8, 1.2)** | **63.3 (7.4, 2.1)** | **64.9 (5.6, 1.0)** | **64.5 (6.1, 0.9)** |
| | | Median (P25, P75) | 63.0 (61.0, 67.8) | 66.3 (63.1, 69.1) | 62.7 (58.5, 69.3) | 65.8 (62.7, 68.4) | 65.4 (61.3, 69.1) |
| | | (Min, Max) | (54.8, 72.8) | (49.04, 72.4) | (47.99, 74.1) | (49.04, 72.8) | (48.0, 74.1) |
| | Age-at-baseline (yrs) | **Mean (SD, SEM)** | **65.4 (5.4, 1.7)** | **66.6 (5.6, 1.2)** | **64.6 (6.5, 1.9)** | **66.2 (5.5, 1.0)** | **65.8 (5.8, 0.9)** |
| | | Median (P25, P75) | 65.0 (63.0, 68.0) | 67.0 (63.0, 71.0) | 66.0 (59.5, 70.0) | 67.0 (63.0, 70.0) | 67.0 (63.0, 70.0) |
| | | (Min, Max) | (55.0, 73.0) | (50.0, 74.0) | (52.0, 74.0) | (50.0, 74.0) | (50.0, 74.0) |
| **Concomitant Medications** | AChE-I | N (%) | 7 (70%) | 13 (56.5%) | 8 (66.7%) | 20 (60.6%) | 28 (62.2%) |
| **Neuroimaging** | Hippocampal vol., % of ICV | **Mean (SD, SEM)** | **0.24 (0.04, 0.01)** | **0.25 (0.04, 0.01)** | **0.28 (0.03, 0.01)** | **0.25 (0.04, 0.01)** | **0.26 (0.04, 0.01)** |
| | | Median (P25, P75) | 0.24 (0.23, 0.27) | 0.25 (0.21, 0.27) | 0.29 (0.27, 0.30) | 0.24 (0.22, 0.27) | 0.26 (0.23, 0.28) |
| | | (Min, Max) | (0.18, 0.29) | (0.17, 0.34) | (0.22, 0.32) | (0.17, 0.34) | (0.17, 0.34) |
| | Ventricular vol., % of ICV | **Mean (SD, SEM)** | **1.84 (0.53, 0.17)** | **3.26 (1.32, 0.28)** | **2.66 (1.08, 0.31)** | **2.83 (1.31, 0.23)** | **2.78 (1.25, 0.19)** |
| | | Median (P25, P75) | 1.81 (1.36, 2.23) | 2.98 (2.07, 4.34) | 2.49 (1.92, 3.29) | 2.65 (1.94, 3.86) | 2.65 (1.93, 3.49) |
| | | (Min, Max) | (1.12, 2.65) | (1.02, 6.20) | (1.07, 5.11) | (1.02, 6.20) | (1.02, 6.20) |
| **CSF Markers** | $A\beta_{42}$, pg/mL | **Mean (SD, SEM)** | **580.7 (109.6, 34.7)** | **700.2 (188.0, 39.2)** | **799.2 (179.6, 51.9)** | **664.0 (175.4, 30.5)** | **700.0 (184.7, 27.5)** |
| | | Median (P25, P75) | 587.5 (479.8, 661.5) | 687.1 (582.7, 849.7) | 823.3 (638.0, 900.1) | 661.5 (563.9, 773.6) | 664.7 (582.7, 845.8) |
| | | (Min, Max) | (428.5, 767.3) | (340.4, 1059.5) | (568.3, 1126.0) | (340.4, 1059.5) | (340.4, 1126.0) |
| | p-$tau_{181}$, pg/mL | **Mean (SD, SEM)** | **35.18 (9.31, 2.94)** | **41.05 (15.24, 3.18)** | **41.27 (13.79, 3.98)** | **39.27 (13.84, 2.41)** | **39.80 (13.70, 2.04)** |
| | | Median (P25, P75) | 32.96 (31.04, 36.07) | 39.36 (28.28, 53.22) | 38.45 (30.01, 51.29) | 36.03 (31.04, 48.31) | 36.07 (30.87, 48.31) |
| | | (Min, Max) | (19.71, 52.59) | (20.55, 81.28) | (23.03, 69.12) | (19.71, 81.28) | (19.71, 81.28) |

| Variable | | | 2 *APOEε4* (N = 10) | 1 *APOEε4* (N = 23) | No *APOEε4* (N = 12) | 1 or 2 *APOEε4* (N = 33) | Overall (N=45) |
|---|---|---|---|---|---|---|---|
| | NfL, pg/mL | **Mean (SD, SEM)** | **1038.51 (319.07, 100.90)** | **1383.13 (438.08, 91.35)** | **1398.91 (296.26, 85.52)** | **1278.70 (431.79, 75.17)** | **1310.75 (400.53, 59.71)** |
| | | Median (P25, P75) | 1074.33 (737.72, 1167.61) | 1395.08 (1091.94, 1572.58) | 1358.55 (1178.45, 1615.49) | 1192.40 (1000.37, 1540.23) | 1296.26 (1042.67, 1540.23) |
| | | (Min, Max) | (630.83, 1718.68) | (580.97, 2391.81) | (999.92, 1992.84) | (580.97, 2391.81) | (580.97, 2391.81) |
| | Ng, pg/mL | **Mean (SD, SEM)** | **481.62 (119.604, 37.82)** | **546.25 (270.94, 56.50)** | **525.67 (235.10, 67.87)** | **526.66 (235.38, 40.97)** | **526.40 (232.62, 34.68)** |
| | | Median (P25, P75) | 472.20 (369.62, 538.99) | 479.52 (307.056, 684.49) | 460.33 (338.81, 696.19) | 474.23 (369.62, 582.30) | 474.23 (362.28, 642.07) |
| | | (Min, Max) | (332.57, 738.87) | (194.62, 1306.92) | (237.78, 1013.23) | (194.62, 1306.92) | (194.62, 1306.92) |
| | YKL-40, ng/mL | **Mean (SD, SEM)** | **201.54 (63.83, 201.86)** | **267.09 (114.89, 239.57)** | **317.58 (161.89, 467.34)** | **247.23 (105.63, 18.39)** | **265.99 (125.13, 18.65)** |
| | | Median (P25, P75) | 223.08 (145.79, 242.01) | 252.31 (197.20, 321.40) | 256.48 (204.27, 433.66) | 235.44 (189.72, 294.80) | 236.94 (197.20, 300.61) |
| | | (Min, Max) | (98.75, 294.80) | (125.21, 598.33) | (131.16, 662.67) | (98.75, 598.33) | (98.75, 662.67) |
| **Cognition** | MMSE Total (0-30) | **Mean (SD, SEM)** | **24.4 (2.1, 0.7)** | **23.3 (2.3, 0.5)** | **23.7 (2.2, 0.6)** | **23.6 (2.3, 0.4)** | **23.6 (2.3, 0.3)** |
| | | Median (P25, P75) | 24.5 (23.0, 26.0) | 23.0 (21.0, 25.0) | 23.5 (21.5, 26.0) | 24.0 (22.0, 26.0) | 24.0 (22.0, 26.0) |
| | | (Min, Max) | (21.0, 27.0) | (20.0, 27.0) | (21.0, 26.0) | (20.0, 27.0) | (20.0, 27.0) |
| | MMSE Memory (0-6) | **Mean (SD, SEM)** | **4.1 (1.0, 0.3)** | **4.0 (1.1, 0.2)** | **4.6 (1.2, 0.4)** | **4.1 (1.1, 0.2)** | **4.2 (1.1, 0.2)** |
| | | Median (P25, P75) | 4.0 (3.0, 5.0) | 4.0 (3.0, 4.0) | 4.5 (3.5, 6.0) | 4.0 (3.0, 4.0) | 4.0 (3.0, 5.0) |
| | | (Min, Max) | (3.0, 6.0) | (2.0, 6.0) | (3.0, 6.0) | (2.0, 6.0) | (2.0, 6.0) |
| | MMSE Visual Construction (0-1) | **Mean (SD, SEM)** | **0.8 (0.4, 0.1)** | **0.7 (0.5, 0.1)** | **0.5 (0.5, 0.2)** | **0.7 (0.5, 0.1)** | **0.6 (0.5, 0.1)** |
| | | Median (P25, P75) | 1.0 (1.0, 1.0) | 1.0 (0.0, 1.0) | 0.5 (0.0, 1.0) | 1.0 (0.0, 1.0) | 1.0 (0.0, 1.0) |
| | | (Min, Max) | (0.0, 1.0) | (0.0, 1.0) | (0.0, 1.0) | (0.0, 1.0) | (0.0, 1.0) |
| | RBANS Total (40-160) | **Mean (SD, SEM)** | **73.4 (10.5, 3.3)** | **64.0 (11.2, 2.3)** | **68.5 (12.3, 3.5)** | **66.8 (11.7, 2.0)** | **67.3 (11.7, 1.7)** |
| | | Median (P25, P75) | 78.0 (61.0, 80.0) | 61.0 (54.0, 73.0) | 67.5 (61.5, 78.5) | 66.0 (57.0, 77.0) | 67.0 (57.0, 77.0) |
| | | (Min, Max) | (57.0, 85.0) | (49.0, 88.0) | (49.0, 90.0) | (49.0, 88.0) | (49.0, 90.0) |
| | | **Mean (SD, SEM)** | **52.8 (12.5, 3.9)** | **48.0 (7.4, 1.6)** | **66.3 (22.5, 6.5)** | **49.5 (9.3, 1.6)** | **53.9 (15.7, 2.3)** |

| Variable | | | 2 *APOEε4* (N = 10) | 1 *APOEε4* (N = 23) | No *APOEε4* (N = 12) | 1 or 2 *APOEε4* (N = 33) | Overall (N=45) |
|---|---|---|---|---|---|---|---|
| | RBANS Delayed Memory (40-154) | Median (P25, P75) | 50.0 (44.0, 56.0) | 48.0 (44.0, 48.0) | 60.0 (48.0, 91.5) | 48.0 (44.0, 52.0) | 48.0 (44.0, 56.0) |
| | | (Min, Max) | (40.0, 84.0) | (40.0, 68.0) | (40.0, 102.0) | (40.0, 84.0) | (40.0, 102.0) |
| | RBANS Visuospatial/ Constructional (40-154) | **Mean (SD, SEM)** | **100.8 (20.0, 6.3)** | **81.1 (19.8, 4.1)** | **79.9 (21.7, 6.3)** | **87.1 (21.6, 3.8)** | **85.2 (21.6, 3.2)** |
| | | Median (P25, P75) | 102.5 (84.0, 121.0) | 78.0 (64.0, 105.0) | 79.5 (61.0, 99.0) | 84.0 (69.0, 105.0) | 81.0 (64.0, 105.0) |
| | | (Min, Max) | (60.0, 121.0) | (50.0, 126.0) | (50.0, 116.0) | (50.0, 126.0) | (50.0, 126.0) |

ICV, intracranial volume; MMSE, Mini-Mental Status Examination; RBANS, Repeatable Battery for the Assessment of Neuropsychological Status; SD, standard deviation; SEM, standard error of the mean.

#### Table S4. Mild AD phenotype in *APOE4* carriers by *APOE4* allele frequency and sex.

| | | | Male | | | Female | | |
|---|---|---|---|---|---|---|---|---|
| | | | **2 *APOEε4* (n = 4)** | **1 *APOEε4* (n = 11)** | ***No APOEε4* (n = 8)** | **2 *APOEε4* (n = 6)** | **1 *APOEε4* (n = 12)** | ***No APOEε4* (n = 4)** |
| **Onset of AD** | Age-at-diagnosis (yrs) | **Mean (SD, SEM)** | **64.4 (5.7, 2.9)** | **65.5 (5.1, 1.5)** | **59.7 (6.1, 2.2)** | **64.0 (5.7, 2.3)** | **65.0 (6.6, 1.90)** | **70.4 (3.2, 1.6)** |
| | | Median (P25, P75) | 62.2 (60.7, 68.1) | 66.3 (63.1, 67.9) | 60.2 (57.3, 62.7) | 64.0 (61.9, 67.8) | 66.1 (63.4, 69.7) | 70.4 (68.0, 72.8) |
| | | (Min, Max) | (60.5, 72.8) | (54.6, 72.4) | (48.0, 69.2) | (54.8, 71.4) | (49.0, 72.1) | (66.6, 74.1) |
| | Age-at-baseline (yrs) | **Mean (SD, SEM)** | **66.5 (4.8, 2.4)** | **67.4 (4.7, 1.4)** | **61.5 (5.5, 1.9)** | **64.7 (6.0, 2.5)** | **65.9 (6.5, 1.9)** | **70.8 (3.4, 1.7)** |
| | | Median (P25, P75) | 65.5 (63.0, 70.0) | 67.0 (63.0, 71.0) | 61.0 (58.5, 66.0) | 64.5 (63.0, 68.0) | 67.5 (63.0, 70.5) | 71.5 (68.5, 73.0) |
| | | (Min, Max) | (62.0, 73.0) | (59.0, 74.0) | (52.0, 69.0) | (55.0, 73.0) | (50.0, 73.0) | (66.0, 74.0) |
| **Concomitant Medications** | AChE-I | N (%) | 3 (75.0%) | 6 (54.5%) | 4 (50.0%) | 4 (66.7%) | 7 (58.3%) | 4 (100.0%) |
| **Neuroimaging** | Hippocampal vol., % of ICV | **Mean (SD, SEM)** | **0.22 (0.03, 0.01)** | **0.24 (0.05, 0.01)** | **0.27 (0.03, 0.01)** | **0.26 (0.03, 0.01)** | **0.25 (0.03, 0.01)** | **0.30 (0.02, 0.01)** |
| | | Median (P25, P75) | 0.23 (0.21, 0.23) | 0.24 (0.19, 0.27) | 0.28 (0.26, 0.29) | 0.27 (0.24, 0.29) | 0.26 (0.23, 0.27) | 0.30 (0.29, 0.32) |
| | | (Min, Max) | (0.18, 0.23) | (0.17, 0.34) | (0.22, 0.30) | (0.21, 0.29) | (0.20, 0.33) | (0.28, 0.32) |
| | Ventricular vol., % of ICV | **Mean (SD, SEM)** | **1.97 (0.28, 0.14)** | **3.53 (1.48, 0.45)** | **2.97 (1.11, 0.39)** | **1.75 (0.66, 0.27)** | **3.01 (1.17, 0.34)** | **2.05 (0.82, 0.41)** |
| | | Median (P25, P75) | 2.04 (1.78, 2.16) | 3.45 (2.09, 4.90) | 3.07 (1.96, 3.40) | 1.49 (1.23, 2.50) | 2.89 (2.00, 3.86) | 2.02 (1.50, 2.59) |
| | | (Min, Max) | (1.57, 2.23) | (1.47, 6.20) | (1.75, 5.11) | (1.12, 2.65) | (1.02, 5.02) | (1.07, 3.06) |
| **CSF Markers** | A$\beta_{42}$, pg/mL | **Mean (SD, SEM)** | **582.1 (139.2, 69.6)** | **699.5 (209.6, 63.2)** | **775.7 (188.9, 66.8)** | **579.7 (100.0, 40.8)** | **700.8 (175.3, 50.6)** | **846.3 (174.9, 87.4)** |
| | | Median (P25, P75) | 545.4 (475.0, 689.2) | 661.6 (491.4, 890.4) | 757.5 (622.9, 875.3) | 606.1 (500.4, 661.5) | 688.0 (612.5, 789.5) | 859.0 (714.5, 978.1) |
| | | (Min, Max) | (470.3, 767.3) | (372.2, 1012.0) | (568.3, 1126.0) | (428.5, 675.8) | (340.4, 1059.5) | (628.3, 1039.0) |
| | p-tau$_{181}$, pg/mL | **Mean (SD, SEM)** | **36.41 (8.33, 4.16)** | **40.05 (14.62, 4.41)** | **35.27 (10.58, 3.74)** | **34.37 (10.60, 4.33)** | **41.96 (16.39, 4.73)** | **53.26 (12.20, 6.10)** |
| | | Median (P25, P75) | 33.53 (30.66, 42.17) | 39.36 (26.50, 54.61) | 33.22 (28.97, 38.45) | 32.96 (31.91, 36.07) | 38.96 (29.70, 52.08) | 51.29 (43.94, 62.58) |

| | | | Male | | | Female | | |
|---|---|---|---|---|---|---|---|---|
| | | | 2 *APOEε4* (n = 4) | 1 *APOEε4* (n = 11) | *No APOEε4* (n = 8) | 2 *APOEε4* (n = 6) | 1 *APOEε4* (n = 12) | *No APOEε4* (n = 4) |
| | | (Min, Max) | (30.29, 48.31) | (20.55, 63.98) | (23.03, 57.86) | (19.71, 52.59) | (23.19, 81.28) | (41.36, 69.12) |
| | NfL, pg/mL | **Mean (SD, SEM)** | **984.23 (215.96, 107.98)** | **1431.56 (346.40, 104.44)** | **1389.31 (346.27, 122.42)** | **1074.70 (389.02, 158.82)** | **1338.73 (519.86, 150.07)** | **1418.09 (203.24, 101.62)** |
| | | Median (P25, P75) | 1015.80 (803.00, 1165.46) | 1420.15 (1348.61, 1622.87) | 1368.45 (1076.94, 1615.49) | 1074.33 (736.10, 1213.90) | 1181.24 (1064.39, 1514.50) | 1358.55 (1285.74, 1550.44) |
| | | (Min, Max) | (737.72, 1167.61) | (807.05, 1903.66) | (999.92, 1992.84) | (630.83, 1718.68) | (580.97, 2391.81) | (1245.68, 1709.57) |
| | Ng, pg/mL | **Mean (SD, SEM)** | **513.22 (158.02, 79.01)** | **557.146 (329.39, 99.32)** | **419.85 (175.80, 62.15)** | **460.54 (97.13, 39.65)** | **536.26 (218.98, 63.22)** | **737.32 (202.38, 101.19)** |
| | | Median (P25, P75) | 472.203 (419.90, 606.55) | 463.83 (271.13, 707.49) | 387.17 (308.79, 460.33) | 483.59 (361.91, 538.99) | 510.607 (366.248, 633.395) | 696.19 ( 592.87, 881.77) |
| | | (Min, Max) | (369.62, 738.87) | (194.62, 1306.92) | (237.78, 808.44) | (332.57, 562.59) | (280.045, 1017.305) | (543.66, 1013.23) |
| | YKL-40, ng/mL | **Mean (SD, SEM)** | **221.97 (23.27, 11.64)** | **260.34 (81.03, 24.43)** | **259.76 (117.50, 41.54)** | **187.93 (80.33, 32.80)** | **273.28 (142.64, 41.18)** | **433.23 (192.70, 96.35)** |
| | | Median (P25, P75) | 228075.3 (205.21, 238.72) | 284.33 (197.20, 321.40) | 237.35 (189.91, 286.86) | 185.62 (112.02, 250.77) | 234.48 (175.04, 312.17) | 433.66 (291.56, 574.90) |
| | | (Min, Max) | (189.72, 242.01) | (134.47, 385.38) | (131.16, 518.66) | (98.75, 294.80) | (125.21, 598.33) | (202.93, 662.67) |
| **Cognition** | MMSE Total (0-30) | **Mean (SD, SEM)** | **25.0 (2.2, 1.1)** | **22.6 (2.5, 0.8)** | **23.4 (2.3, 0.8)** | **24.0 (2.2, 0.9)** | **23.8 (2.1, 0.6)** | **24.3 (2.4, 1.2)** |
| | | Median (P25, P75) | 25.5 (23.5, 26.5) | 22.0 (20.0, 26.0) | 22.5 (21.5, 26.0) | 23.5 (23.0, 26.0) | 24.5 (22.5, 25.0) | 25.0 (22.5, 26.0) |
| | | (Min, Max) | (22.0, 27.0) | (20.0, 26.0) | (21.0, 26.0) | (21.0, 27.0) | (20.0, 27.0) | (21.0, 26.0) |
| | MMSE Memory (0-6) | **Mean (SD, SEM)** | **4.0 (0.8, 0.4)** | **3.8 (0.9, 0.3)** | **4.6 (1.3, 0.5)** | **4.2 (1.2, 0.5)** | **4.3 (1.3, 0.4)** | **4.5 (1.3, 0.6)** |
| | | Median (P25, P75) | 4.0 (3.5, 4.5) | 4.0 (3.0, 4.0) | 4.5 (3.5, 6.0) | 4.0 (3.0, 5.0) | 4.0 (3.5, 5.5) | 4.5 (3.5, 5.5) |
| | | (Min, Max) | (3.0, 5.0) | (3.0, 6.0) | (3.0, 6.0) | (3.0, 6.0) | (2.0, 6.0) | (3.0, 6.0) |
| | MMSE Visual Construction (0-1) | **Mean (SD, SEM)** | **1.0 (0.0, 0.0)** | **0.5 (0.5, 0.2)** | **0.3 (0.5, 0.2)** | **0.7 (0.5, 0.2)** | **0.8 (0.4, 0.1)** | **1.0 (0.0, 0.0)** |
| | | Median (P25, P75) | 1.0 (1.0, 1.0) | 0.0 (0.0, 1.0) | 0.0 (0.0, 0.5) | 1.0 (0.0, 1.0) | 1.0 (1.0, 1.0) | 1.0 (1.0, 1.0) |
| | | (Min, Max) | (1.0, 1.0) | (0.0, 1.0) | (0.0, 1.0) | (0.0, 1.0) | (0.0, 1.0) | (1.0, 1.0) |

| | | Male | | | Female | | |
|---|---|---|---|---|---|---|---|
| | | **2 *APOEε4* (n = 4)** | **1 *APOEε4* (n = 11)** | ***No APOEε4* (n = 8)** | **2 *APOEε4* (n = 6)** | **1 *APOEε4* (n = 12)** | ***No APOEε4* (n = 4)** |
| MMSE Attention/Calculation (0-5) | **Mean (SD, SEM)** | **3.5 (1.9, 1.0)** | **3.5 (1.3, 0.4)** | **2.8 (1.7, 0.6)** | **3.5 (1.5, 0.6)** | **3.8 (1.5, 0.4)** | **3.8 (1.0, 0.5)** |
| | Median (P25, P75) | 4.0 (2.0, 5.0) | 4.0 (3.0, 5.0) | 3.0 (1.0, 4.0) | 3.5 (3.0, 5.0) | 4.0 (3.5, 5.0) | 3.5 (3.0, 4.5) |
| | (Min, Max) | (1.0, 5.0) | (1.0, 5.0) | (1.0, 5.0) | (1.0, 5.0) | (1.0, 5.0) | (3.0, 5.0) |
| RBANS Total (40-160) | **Mean (SD, SEM)** | **74.0 (11.6, 5.8)** | **61.6 (9.2, 2.8)** | **67.9 (14.8, 5.2)** | **73.0 (10.8, 4.4)** | **66.1 (12.7, 3.7)** | **69.8 (6.3, 3.2)** |
| | Median (P25, P75) | 78.0 (67.0, 81.0) | 60.0 (54.0, 71.0) | 64.5 (56.5, 81.0) | 76.5 (61.0, 80.0) | 65.0 (54.0, 76.5) | 70.0 (65.0, 74.5) |
| | (Min, Max) | (57.0, 83.0) | (49.0, 76.0) | (49.0, 90.0) | (59.0, 85.0) | (49.0, 88.0) | (62.0, 77.0) |
| RBANS Delayed Memory (40-154) | **Mean (SD, SEM)** | **48.0 (7.3, 3.7)** | **46.9 (6.9, 2.1)** | **67.9 (25.0, 8.8)** | **56.0 (14.8, 6.0)** | **49.0 (8.0, 2.3)** | **63.0 (19.7, 9.8)** |
| | Median (P25, P75) | 48.0 (42.0, 54.0) | 48.0 (40.0, 48.0) | 64.0 (44.0, 92.5) | 50.0 (48.0, 60.0) | 48.0 (44.0, 50.0) | 56.0 (52.0, 74.0) |
| | (Min, Max) | (40.0, 56.0) | (40.0, 64.0) | (40.0, 102.0) | (44.0, 84.0) | (40.0, 68.0) | (48.0, 92.0) |
| RBANS Visuospatial/ Constructional (40-154) | **Mean (SD, SEM)** | **104.3 (16.0, 8.0)** | **78.2 (23.7, 7.1)** | **70.9 (21.0, 7.4)** | **98.5 (23.4, 9.6)** | **83.8 (16.2, 4.7)** | **98.0 (7.1, 3.5)** |
| | Median (P25, P75) | 106.0 (92.0, 116.5) | 64.0 (64.0, 105.0) | 63.0 (58.0, 79.5) | 102.5 (84.0, 121.0) | 78.0 (72.0, 94.5) | 99.0 (92.5, 103.5) |
| | (Min, Max) | (84.0, 121.0) | (50.0, 126.0) | (50.0, 116.0) | (60.0, 121.0) | (64.0, 112.0) | (89.0, 105.0) |
| RBANS Attention (40-154) | **Mean (SD, SEM)** | **82.8 (13.0, 6.5)** | **73.7 (12.9, 3.9)** | **71.4 (9.4, 3.3)** | **74.8 (24.8, 10.1)** | **79.8 (15.2, 4.4)** | **81.0 (14.2, 7.1)** |
| | Median (P25, P75) | 86.5 (74.5, 91.0) | 72.0 (64.0, 79.0) | 73.5 (68.0, 75.0) | 73.5 (53.0, 82.0) | 83.5 (68.0, 91.0) | 86.5 (72.5, 89.5) |
| | (Min, Max) | (64.0, 94.0) | (56.0, 106.0) | (53.0, 85.0) | (49.0, 118.0) | (53.0, 100.0) | (60.0, 91.0) |

ICV, intracranial volume; MMSE, Mini-Mental Status Examination; RBANS, Repeatable Battery for the Assessment of Neuropsychological Status; SD, standard deviation; SEM, standard error of the mean.
